# Reverse cholesterol transport and lipid peroxidation biomarkers in major depression and bipolar disorder: a systematic review and meta-analysis

**DOI:** 10.1101/2023.03.20.23287483

**Authors:** Abbas F. Almulla, Yanin Thipakorn, Ali Abbas Abo Algon, Chavit Tunvirachaisakul, Hussein K. Al-Hakeim, Michael Maes

## Abstract

**Background:** Major depression (MDD) and bipolar disorder (BD) are linked to immune activation, increased oxidative stress, and lower antioxidant defenses.

**Objectives:** To systematically review and meta-analyze all data concerning biomarkers of reverse cholesterol transport (RCT), lipid-associated antioxidants, lipid peroxidation products, and autoimmune responses to oxidatively modified lipid epitopes in MDD and BD.

**Methods:** Databases including PubMed, Google scholar and SciFinder were searched to identify eligible studies from inception to January 10th, 2023. Guidelines of Preferred Reporting Items for Systematic Reviews and Meta-Analyses (PRISMA) guidelines were followed.

**Results:** The current meta-analysis included 176 studies (60 BD and 116 MDD) and examined 34,051 participants, namely 17,094 with affective disorders and 16,957 healthy controls. Patients with MDD and BD showed a) significantly decreases in RCT (mainly lowered high-density lipoprotein cholesterol and paraoxonase 1); b) lowered lipid soluble vitamins (including vitamin A, D, and coenzyme Q10); c) increased lipid peroxidation and aldehyde formation, mainly increased malondialdehyde (MDA), 4-hydroxynonenal, peroxides, and 8-isoprostanes; and d) Immunoglobulin (Ig)G responses to oxidized low-density lipoprotein and IgM responses to MDA. The ratio of all lipid peroxidation biomarkers / all lipid-associated antioxidant defenses was significantly increased in MDD (standardized mean difference or SMD=0.433; 95% confidence intervals (CI): 0.312; 0.554) and BD (SMD=0.653; CI: 0.501-0.806). This ratio was significantly greater in BD than MDD (p=0.027).

**Conclusion:** In MDD/BD, lowered RCT, a key antioxidant and anti-inflammatory pathway, may drive increased lipid peroxidation, aldehyde formation, and autoimmune responses to oxidative specific epitopes, which all together cause increased immune-inflammatory responses and neurotoxicity.

## Introduction

There is now evidence that major depression (MDD) and bipolar disorder (BD) are immune disorders characterized by activation of the immune-inflammatory response system (IRS) and the compensatory immunoregulatory system (CIRS) (Almulla and Maes, 2022; Maes, 1995; Maes and Carvalho, 2018; Maes et al., 2011c; Vasupanrajit et al., 2022). IRS activation in MDD is indicated by increased levels of key cytokines of the M1 macrophage, T-helper (Th)-1, Th-2, Th-17, and T regulatory (Treg) lineages, as well as increased T cell activation markers on peripheral blood mononuclear cells (Maes and Carvalho, 2018; Maes et al., 1992).

Most immune functions are mediated and regulated via redox mechanisms, including the regulatory effects of antioxidant defenses (Morris et al., 2022). Moreover, immune activation is accompanied by increased concentrations of reactive oxygen (ROS) and nitrogen (RNS) species, including superoxide, peroxides, nitric oxide (NO), and peroxynitrite leading to nitro-oxidative damage to lipids, proteins and DNA, which may aggravate immune responses (Maes et al., 2011a; Moylan et al., 2014). Furthermore, lowered levels of antioxidants predispose towards nitro-oxidative stress and inflammatory responses (Maes et al., 2011a). In MDD and BD, activated neuro-oxidative pathways are associated with aberrations in neuro-immune pathways (Maes et al., 2011a; Moylan et al., 2014; Sowa-Kućma et al., 2018).

The first publication on deficits in lipid-associated antioxidant defenses was published in 1994 (Maes et al., 1994), reporting impairments in reverse cholesterol transport (RCT). The latter is a major detoxification pathway which removes free cholesterol from the body, thereby preventing atherogenicity and protecting against lipid peroxidation, oxidation of low (LDL) and high density lipoprotein (HDL) cholesterol and inflammatory responses (Jirakran et al., 2023; Maes et al., 1994; Maes et al., 1997). Deficits in RCT in MDD, as indicated by lowered HDL, apolipoprotein A1 (ApoA) and lecithin-cholesterol acyltransferase (LCAT) activity, and increased free cholesterol levels (Jirakran et al., 2023; Maes et al., 1994; Maes et al., 1997), may cause increased atherogenicity (Maes et al., 1994), as well as damage to lipids with formation of oxidized LDL (oxLDL) and HDL (oxHDL) and formation of aldehydes, including malondialdehyde (MDA) (Maes et al., 2011e). Moreover, neoepitopes such as oxLDL and MDA are immunogenic and may activate the IRS and cause immunoglobulin (Ig)G and IgA-mediated autoimmune responses (Maes et al., 2011d). Furthermore, MDD is not only accompanied by lowered RCT, HDL, ApoA and LCAT, but also by lowered paraoxonase 1 (PON1) enzyme activity (Moreira et al., 2019b; Moreira et al., 2019a). This is important because PON1 has antioxidant, anti-inflammatory, and antiatherogenic activities (Moreira et al., 2019b; Moreira et al., 2019a). Apo A, LCAT and PON1 are part of the HDL complex and determine the antioxidant capacity of the HDL particles (Jirakran et al., 2023; Moreira et al., 2019b; Moreira et al., 2019a).

High concentrations of ROS/RNS and IRS activation may impact antioxidant defenses for example, by inducing lowered levels of circulating HDL (Feingold and Grunfeld, 2016; Guirgis et al., 2018; Kim et al., 2020), ApoA1 (Guirgis et al., 2018; Smith, 2010; Undurti et al., 2009) and PON1 (Han et al., 2006; Kumon et al., 2003; Moreira et al., 2019b; Moreira et al., 2019a). During inflammatory conditions, increased activity of myeloperoxidase (MPO) may impair PON1 activity and, thus, the antioxidant protection of HDL, which eventually may become pro-inflammatory (Farid and Horii, 2012; Han et al., 2006; Kumon et al., 2003; Morris et al., 2022). As such, MDD is characterized by intertwined aberrations in IRS and lipid peroxidation and lowered lipid-associated antioxidant defenses (Maes et al., 2011b).

Dysfunctions in the HDL-PON1-ApoA-LCAT complex and RCT are considered to be key factors in affective disorders. This is because the Q192R PON1 variant, which controls PON1 activity and the antioxidant properties of the HDL-complex, is strongly linked to MDD and BD (Jirakran et al., 2023; Maes et al., 2011e; Morris et al., 2022). As such, PON1 gene variants, which determine lowered PON1 activity and, thus, lowered activity of the HDL-PON1-ApoA-LCAT complex predispose to increased lipid peroxidation, autoimmune responses, immune activation and increased atherogenicity (Jirakran et al., 2023; Maes et al., 1997). This is important because the HDL-PON1 and HDL-ApoA complexes are strongly associated with the recurrence of illness (ROI) of MDD and BD, suicidal behaviors and severity of depression (Jirakran et al., 2023; Maes et al., 2021; Maes et al., 2019; Moreira et al., 2019b; Moreira et al., 2019a; Vasupanrajit et al., 2022). Moreover, ApoE and ghrelin are endogenous antioxidants that protect against inflammation and take part in the HDL-PON1-ApoA-LCAT complex (Akki et al., 2021; Daniil et al., 2011).

Clinical studies and animal models of depression reported high oxidative products, including biomarkers of lipid peroxidation, such as MDA and 8-iso-prostaglandin F2 (8-iso) (Dimopoulos et al., 2008; Gałecki et al., 2009; Yager et al., 2010). Also, lowered levels of fat-soluble antioxidants (vitamins A, D, E, and K) (Khanzode et al., 2003; Maes et al., 2000b; Owen et al., 2005), and antioxidants that protect against lipid peroxidation, including coenzyme Q10, are frequently observed in MDD and BD (Maes et al., 2009; Morris et al., 2021b). A first meta-analysis study showed that depression is accompanied by increased red-blood cell and serum levels of MDA, peroxides, 8-F2-isoprostanes and decreased levels of antioxidants including PON1 and HDL, and antioxidants that protect against ROS production, including albumin and zinc (Liu et al., 2015; Mazereeuw et al., 2015; Wei et al., 2020). Other meta-analyses showed that BD and MDD are characterized by increased lipid peroxidation and attenuated antioxidant defenses (Black et al., 2015; Brown et al., 2014; Jiménez-Fernández et al., 2021).

Nonetheless, these meta-analyses did not examine the more comprehensive lipid peroxidation profiles, including evaluation of the HDL-PON1-ApoA-LCAT complex (RCT), ghrelin and Apo E, and other lipid-associated antioxidants (LPANTIOX), including (vitamins A, D, E, K and coenzyme Q10; labeled ADEKQ) versus lipid associated oxidative stress toxicity (LPOSTOX), including assessments of lipid hydroperoxides (LOOH), MDA, 8-isoprostane, 4-hydroxynonenal (4-HNE), oxLDL and autoimmune responses to neoepitopes including MDA and oxLDL (LPAUTO). We also aimed to examine the LPOSTOX + LPAUTO / ADECKQ + RCT (together labeled LPANTIOX) ratio to examine the net effect of detrimental versus protective redox-related factors in MDD/BD as well as difference between MDD and BD.

## Materials and method

We adhered to various methodological guidelines to conduct the current study, namely Preferred Reporting Items for Systematic Reviews and Meta-Analyses (PRISMA) 2020 (Page et al., 2021), Cochrane Handbook for Systematic Reviews and Interventions (Cumpston et al., 2019), and the Meta-Analyses of Observational Studies in Epidemiology (MOOSE). We recruited MDD and BD patients in the current meta-analysis to examine central and peripheral levels of LPOSTOX biomarkers, namely LOOH, MDA/Thiobarbituric acid reactive substances (TBARS), 4-HNE, isoprostanes; LPAUTO biomarkers including IgM/IgG responses to neoepitopes including MDA, and oxLDL, RCT biomarkers, including (HDL, PON1, LCAT, ApoA), and ADEKQ vitamins, including CoQ10). Consequently, based on the aforementioned and other biomarkers (see below), we computed composites reflecting LPOSTOX, LPAUTO, RCT, ADECQ, and LPANTIOX.

### Search strategy

To gather all relevant data on the biomarkers, we searched electronic databases PubMed/MEDLINE, Google Scholar, and SciFinder using prespecified keywords and Mesh terms outlined in Table 1 of the supplementary electronic file (ESF), from September to January 10th, 2023. Additionally, we inspected the reference lists of eligible studies and preceding meta-analyses to ensure no pertinent studies were missed.

**Table 1.** The outcomes and number of patients with affective disorders and healthy controls and the position of the standardized mean difference (SMD) and 95% confidence intervals with respect to the zero SMD.

| Outcome profiles | n studies | Side of 95% confidence intervals |  |  |  | Patient Cases | Control Cases | Total number of participants |
| --- | --- | --- | --- | --- | --- | --- | --- | --- |
|  |  | < 0 | Overlap 0 and SMD < 0 | Overlap 0 and SMD > 0 | > 0 |  |  |  |
| RCT | 55 | 18 | 19 | 13 | 5 | 10372 | 11073 | 21445 |
| ADECK | 27 | 16 | 6 | 4 | 1 | 2690 | 2045 | 4735 |
| LPANTIOX | 99 | 39 | 23 | 22 | 15 | 13541 | 13692 | 27233 |
| LPOSTOX | 93 | 4 | 11 | 21 | 55 | 4538 | 4189 | 8727 |
| LPAUTO | 7 | 0 | 0 | 0 | 7 | 394 | 230 | 624 |
| (LPOSTOX+LPAUTO) / LPANTIOX | 176 | 16 | 27 | 44 | 89 | 17106 | 16921 | 34027 |

RCT or reverse cholesterol transport comprises high-density lipoprotein (HDL), paraoxonase-1 (PON-1), apolipoprotein A (Apo A), lecithin cholesterol acyltransferase (LCAT), ApoE and ghrelin.
ADECK comprises vitamin A, D, E, K, and coenzyme Q10.
LPANTIOX or lipid-associated antioxidant defenses comprises RCT and ADECK.
LPOSTOX or lipid oxidative stress toxicity comprises MDA, TBARS, 8-Isoprostane, 4-HNE, oxLDL, 7-Ketocholesterol, 27-Hydroxycholesterol, 25-Hydroxycholesterol, 24-Hydroxycholesterol, peroxides, LOOH and ApoB.
LPAUTO or autoimmunity against lipid associated neopeptides. comprises immunoglobulin G against oxLDL and Immunoglobulin M against MDA.

### Eligibility criteria

The main criteria for including articles in this meta-analysis were that they were published in peer-reviewed journals and written in English. Nevertheless, the study also took into consideration grey literature and articles written in various languages, including Thai, French, Spanish, Turkish, German, Italian, and Arabic. Furthermore, inclusion criteria were observational case-control and cohort studies, which include controls and assessed lipid peroxidation and lipid antioxidant biomarkers. The MDD and BD patients should have been diagnosed according to the Diagnostic and Statistical Manual of Mental Disorders (DSM) or International Classification of Diseases (ICD). Follow up studies could be included provided that baseline levels of the biomarkers were assessed. Exclusion criteria were: a) animal-based, genotypic and translational studies, b) studies without a control group, c) studies with uncommon media, namely saliva, hair, whole blood, and platelet-rich plasma, d) papers that did not provide the mean and standard deviation (SD) or standard error (SE) values of the evaluated biomarkers. However, we sent emails to the authors of studies lacking mean (SD) or (SE) values and asked the authors to provide mean and SD values. In case of no response, we computed the mean (SD) values from the median (interquartile, IQR or minimum-maximum, min-mix range) based on the approach proposed by Wan et al (Wan et al., 2014) and estimated the mean (SD) of graphical data utilizing the Web Plot Digitizer (https://automeris.io/WebPlotDigitizer/).

### Primary and secondary outcomes

The primary outcomes of the present meta-analysis are the RCT, ADECQ, LPANTIOX, LPOSTOX, and LPAUTO biomarker profiles as shown in **Table 1**. Subsequently, if these composites were significant, we also examined their single indicators.

### Screening and data extraction

Two authors, AA and YT, carried out a preliminary evaluation of the relevant studies for inclusion in the meta-analysis. This evaluation was performed by reviewing the titles and abstracts of the studies and applying the predetermined inclusion criteria. Subsequently, we obtained full texts of the papers that met our inclusion criteria and disregarded those that did meet our exclusion criteria. The same two authors employed a pre-specified excel file that included data like author names, study dates, biomarker names, mean and SD of the biomarkers, number of patients and control groups, study sample sizes. Furthermore, we also included the study design, type of sample (serum, plasma, CSF, brain tissues, blood cells), psychiatric rating scales used, mean (SD) of participants’ ages, sex and location of the study. The authors double-checked the file and resorted to the last author (MM) in case of discrepancies.

Additionally, the quality of the methodology of the included studies was assessed utilizing the Immunological Confounder Scale (ICS) (Andres-Rodriguez et al., 2020). The ICS was adjusted by the last author (MM) to make it in line with lipid peroxidation studies. The ICS comprises two evaluation scales, namely the Quality Scale and the Redpoints Scale, which are outlined in Table 2 of the ESF. These scales were used in previous meta-analysis to gauge the methodological rigor of the publications that examined the immune and tryptophan catabolite data in individuals with affective disorders (Almulla et al., 2022b; Almulla et al., 2022c; Vasupanrajit et al., 2022). The Quality Scale score ranges from 0 to 10, where 0 represents lower quality and 10 indicates better quality. The scale mainly evaluates factors such as sample size, control of confounding variables and sampling duration. The redpoints scale is primarily designed to detect potential bias in the lipid peroxidation results and research designs by assessing the level of control over critical confounding factors. When the overall score was 0, it indicates that the highest level of control was maintained. Conversely, a score of 26 indicates that there was no control for the confounding variables.

**Table 2.** Results of meta-analyses performed on the primary outcome variables.

| Outcome feature sets | N | Groups | SMD | 95% CI | z | p | Q | df | p | I<sup>2</sup> (%) | $\tau^2$ | T |
| --- | --- | --- | --- | --- | --- | --- | --- | --- | --- | --- | --- | --- |
| RCT* | 55 | Overall | -0.286 | -0.403; -0.169 | -4.804 | 0.000 | 527.277 | 54 | <0.0001 | 89.759 | 0.150 | 0.387 |
| ADECK* | 27 | Overall | -0.533 | -0.763; -0.304 | -4.557 | <0.0001 | 322.595 | 26 | <0.0001 | 91.940 | 0.317 | 0.563 |
| LPANTIOX* | 99 | Overall | -0.270 | -0.374; -0.165 | -5.053 | <0.0001 | 1175.144 | 98 | <0.0001 | 91.661 | 0.226 | 0.475 |
| LPOSTOX* | 93 | Overall | 0.731 | 0.572; 0.891 | 8.982 | <0.0001 | 1070.419 | 92 | <0.0001 | 91.405 | 0.542 | 0.736 |
| LPAUTO* | 7 | Overall | 0.904 | 0.727; 1.081 | 10.011 | <0.0001 | 5.448 | 6 | 0.488 | 0.000 | 0.000 | 0.000 |
| (LPOSTOX+LPAUTO)/<br>LPANTIOX | 60 | BD | 0.653 | 0.501; 0.806 | 8.383 | 0.000 | 604.682 | 59 | 0.000 | 90.243 | 0.291 | 0.540 |
|  | 116 | Depression | 0.433 | 0.312; 0.554 | 7.007 | 0.000 | 1441.883 | 115 | 0.000 | 92.024 | 0.380 | 0.617 |
|  | **BD Vs depression Q= 4.918, df=1, p= 0.027** |  |  |  |  |  |  |  |  |  |  |  |

\* Not significantly different between bipolar disorder and major depression
RCT or reverse cholesterol transport comprises high-density lipoprotein (HDL), paraoxonase-1 (PON-1), apolipoprotein A (ApoA), lecithin cholesterol acyltransferase (LCAT), ApoE and ghrelin.
ADECK comprises vitamin A, D, E, K, and coenzyme Q10.
LPANTIOX or lipid-associated antioxidant defenses comprises RCT and ADECK.
LPOSTOX or lipid oxidative stress toxicity comprises MDA, TBARS, 8-Isoprostane, 4-HNE, oxLDL, 7-Ketocholesterol, 27-Hydroxycholesterol, 25-Hydroxycholesterol, 24-Hydroxycholesterol, peroxides, LOOH and ApoB.
LPAUTO or autoimmunity against lipid associated neopeptides. comprises immunoglobulin G against oxLDL and Immunoglobulin M against MDA.

### Data analysis

We employed CMA V4 software while following PRISMA guidelines (see ESF, Table 3) to conduct the current meta-analysis, which required a minimum of two studies on each determined biomarker. LPOXTOX was computed by assuming dependence and including all relevant biomarkers in the CMA (MDA, TBARS, isoprostanes, 4-HNE, oxidized LDL, peroxide, LOOH, hydroxy and keto cholesterol, Apo B). LPAUTO included IgG responses to oxLDL and IgM responses to neoepitopes, including MDA. Likewise, we computed RCT by including PON1, LCAT, HDL and ApoA, ApoE and ghrelin, ADEKQ including vitamin A, D, E and K, and CoQ10, and LPANTIOX (RCT and ADECK). Furthermore, in the present meta-analysis, we also computed (LPOSTOX + LPAUTO) / LPANTIOX ratio by selecting a positive effect size for all LPOSTOX and LPAUTO data and a negative effect size for all LPANTIOX biomarkers in patients versus healthy controls while assuming dependence. We employed a random-effect model with constrained maximum likelihood to pool the effect sizes. Statistical significance was defined as a two-tailed p-value less than 0.05, and effect sizes are reported as the standardized mean difference (SMD) with 95% confidence intervals (95% CI). SMD of 0.80, 0.5, and 0.20 were used to classify the effect size as large, moderate, or small, respectively (Cohen, 2013). We assessed heterogeneity by computing tau-squared statistics besides the Q and I^2^ values as previously used in meta-analyses (Almulla et al., 2022a; Almulla et al., 2022d; Vasupanrajit et al., 2022). In addition, we used meta-regression to delineate the factors contributing to the observed heterogeneity. The divergences in biomarkers among MDD and BD patients and between various media types, brain tissues and cerebrospinal fluid (CSF), serum, plasma and blood cells, were examined by subgroup analysis with each of the determinants being used as a unit of analysis. CSF and brain tissues were grouped together under the central nervous system (CNS) category due to the absence of statistically significant differences between studies measuring the biomarkers in these compartments. Likewise, we combined the studies of MDD and BD that measured lipid peroxides and antioxidants; however, we present the effect sizes of MDD and BD separately when there was a significant difference.

**Table 3.**
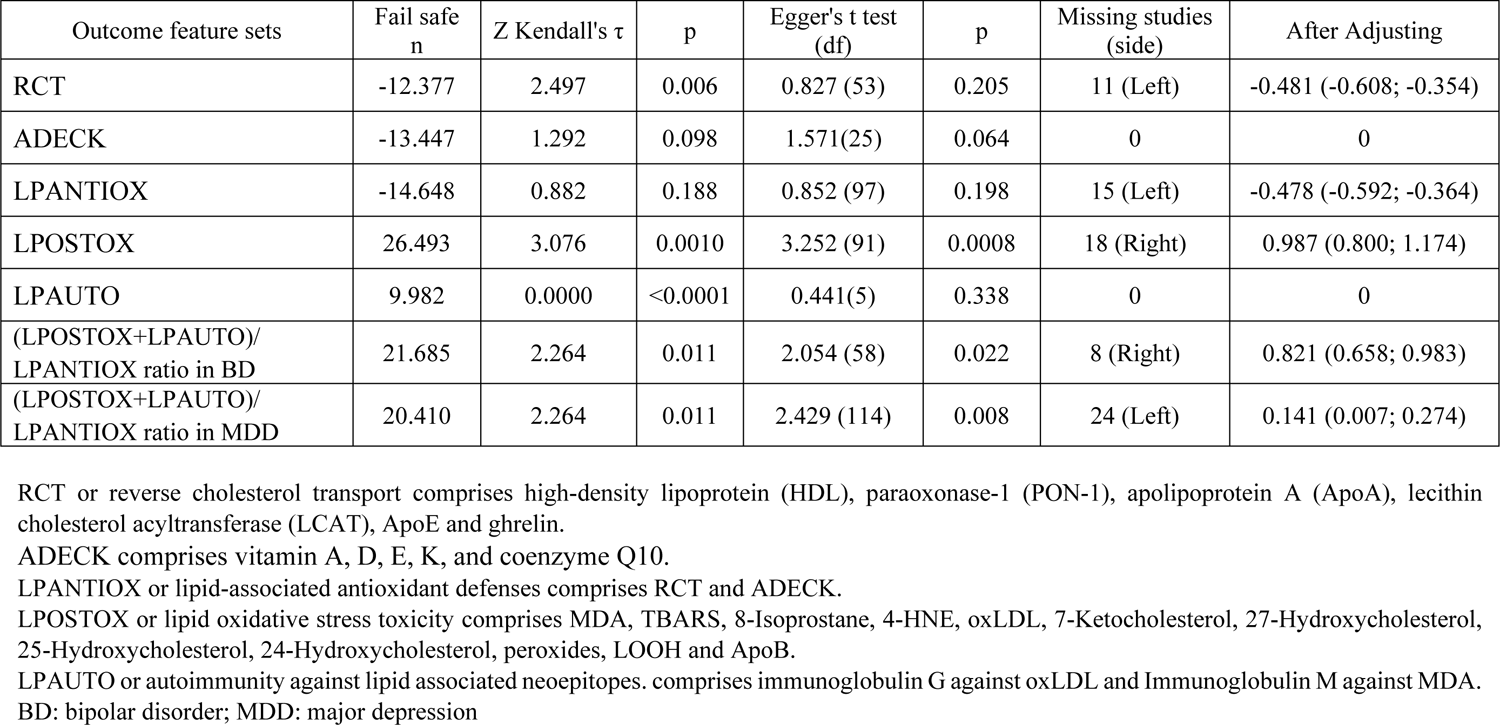
Results on publication bias.

To evaluate the stability of effect sizes, we performed sensitivity analyses using the leave-one-out method. We examined the presence of publication bias by utilizing the fail-safe N technique, the continuity-corrected Kendall tau, and the Egger’s regression intercept, with one-tailed p-values being used for the latter two methods. The trim-and-fill approach was used to impute the missing studies in case of an asymmetry indicated by Egger’s test; and adjusted effect sizes were computed. We also utilized funnel plots (study precision vs SMD), which simultaneously display observed and imputed missing values, to identify small study effects.

## Results Search results

In the initial search process, we found 56157 studies based on our predefined particular keywords and Mesh terms (see ESF, Table 1). The PRISMA flow chart in **Figure 1** illustrates the search results, including the total number of included and excluded articles. Nevertheless, 55136 articles were excluded after the results had been refined and irrelevant studies were removed. The current systematic review involved 169 studies, as 852 articles were removed due to our inclusion-exclusion criteria. Moreover, 6 studies were eliminated due to reasons mentioned in ESF Table 4. As a consequence, 163 studies were eligible based on inclusion-exclusion criteria and entered in the present meta-analysis (see ESF, Eligible studies).

**Figure 1:**
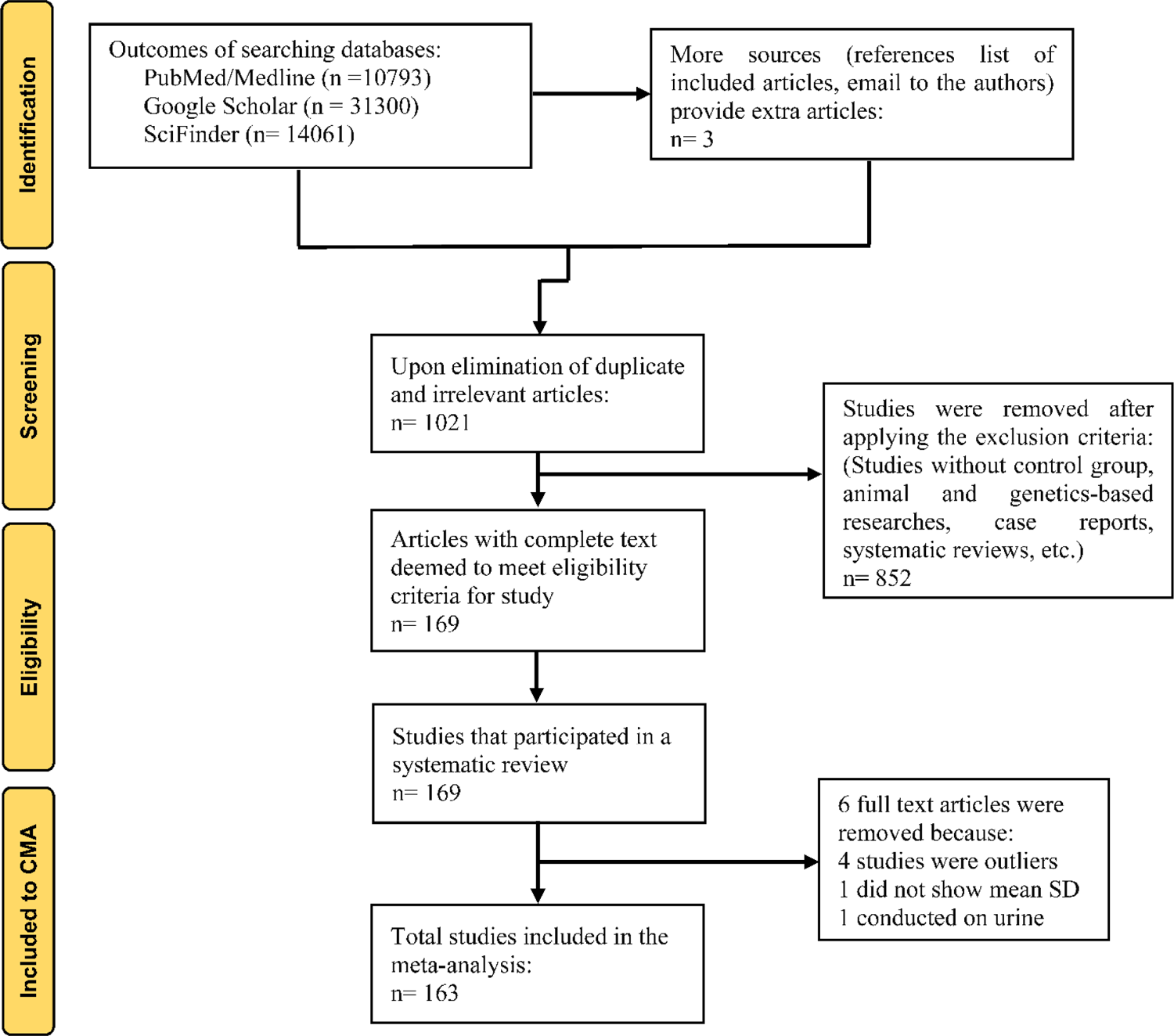
The PRISMA flow chart.

Furthermore, fourteen of the included articles examined lipid peroxidation biomarkers in two cohorts of patients, MDD and BD, within the same research and 1 study examined central and peripheral biomarkers. Consequently, the overall effect size in the current meta-analysis was pooled from 176 studies (60 BD and 116 MDD). We employed totally 34051 participants, namely 17094 affective disorder patients and 16957 healthy controls. The age ranged from 26 – 65 years. Several techniques were employed to assess the biomarkers examined in the current meta-analysis with ELISA; spectrophotometric and automated analysis being the most used techniques (ESF, Table 5). Turkey and Brazil have contributed the most to the eligible research, with 38 and 21 publications, respectively, while numerous other countries have contributed between 1 and 9 studies. In the present meta-analysis, the quality and redpoint scores expressed as median (min-max) were computed, and the findings are given in ESF, table 5.

### Primary outcome variables RCT profile

**Table 1** shows that our meta-analysis comprises 55 studies that were analyzed to investigate the composite score of RCT. Of these studies, 18 displayed confidence intervals (CIs) that were entirely below zero, while 5 had CIs that was entirely above zero. In addition, 32 studies demonstrated overlapping CIs, and the SMD values were negative in 19 studies and positive in 13, as shown in Table 1. The forest plot in **Figure 2** depicts that MDD and BD patients exhibit significantly reduced RCT levels (effect size of −0.286) compared to healthy controls. Publication bias analysis revealed 11 studies missing to the left of the funnel plot. Imputing these studies led to a change in the SMD value, which became −0.481.

**Figure 2:**
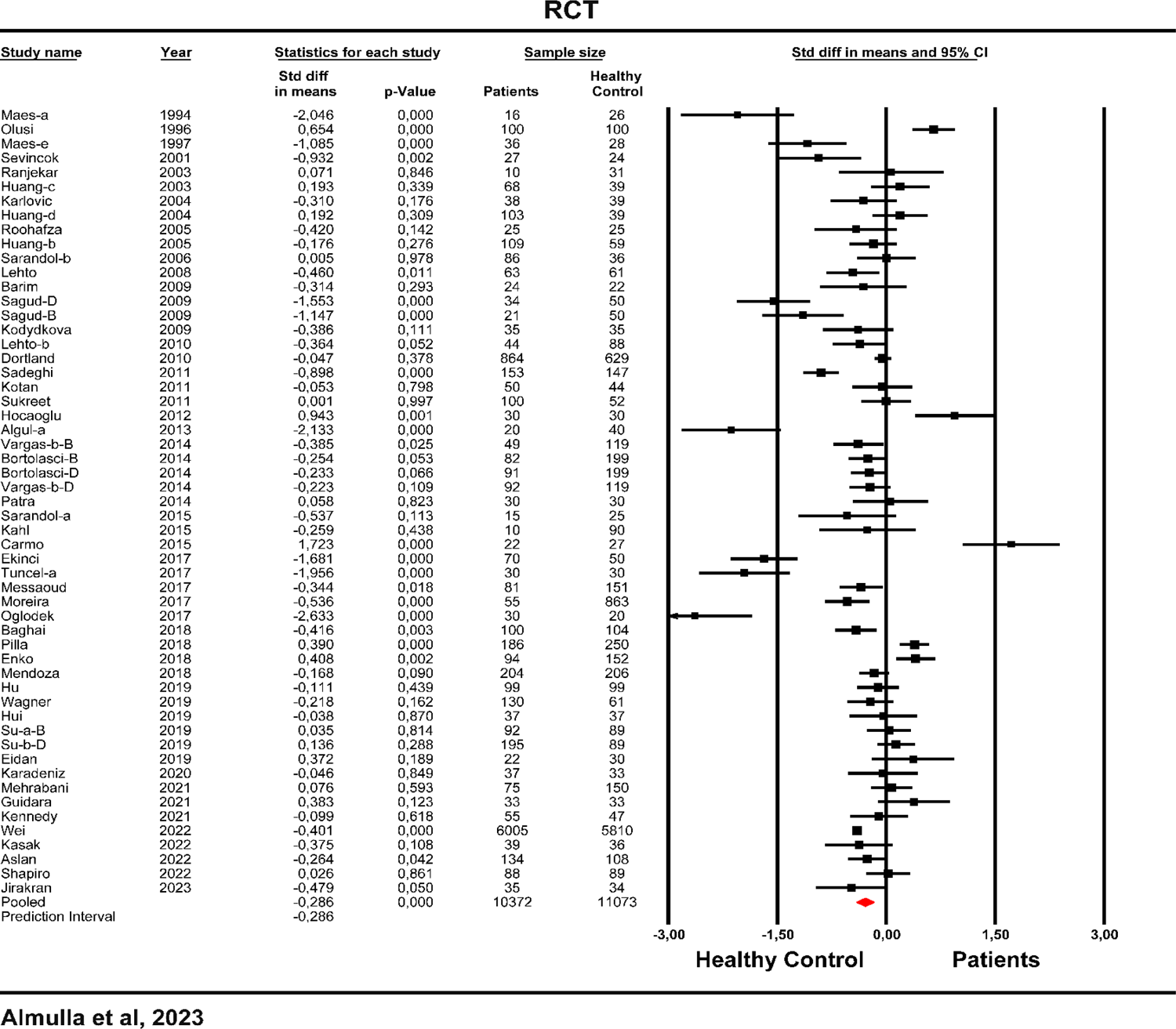
Forest plot of reverse cholesterol transport (RCT) in patients with major depression and bipolar disorder compared to healthy controls.

### ADECQ profile (lipid soluble vitamins and Coenzyme Q10)

In the present meta-analysis, we incorporated 27 relevant studies to investigate the ADECQ index in MDD and BD patients. **Table 1** illustrates that out of these studies, 16 had CIs that were entirely negative, while only one study had a CI that was entirely positive. Furthermore, zero-overlapping CIs were observed in 10 studies, and among them, the SMD values were less than zero in 6 studies and greater than zero in 4 studies. **Table 2** and **Figure 3** indicate that MDD and BD patients display a significant decrease with a moderate effect size (−0.533) in the ADECQ profile. **Table 3** demonstrates that no bias was detected.

**Figure 3:**
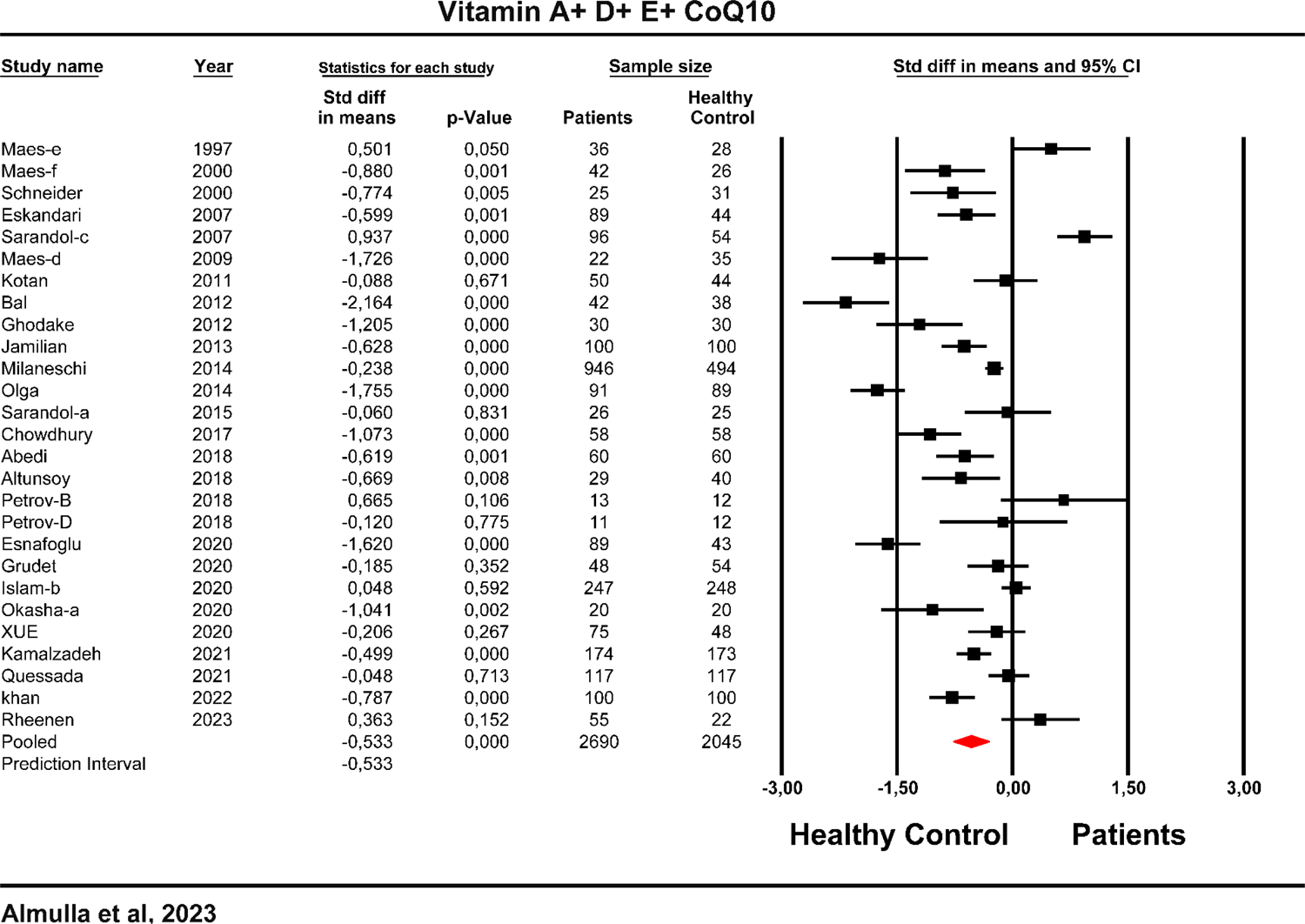
Forest plot of vitamins A, D, and E and coenzyme Q10 in patients with major depression and bipolar disorder as compared to healthy controls.

### LPANTIOX profile

The present study investigated the LPANTIOX composite score by integrating data from 99 studies. **Table 1** demonstrate that 39 studies yielded CIs that were smaller than zero, whereas 15 studies yielded CIs that were greater than zero. **Table 2** and **Figure 4** reveal that patients with MDD and BD, as compared to healthy controls, exhibit significantly reduced lipid-associated antioxidant defenses, as indicated by a lowered LPANTIOX composite score (SMD = −0.270). Egger’s and Kendall’s tests revealed a significant bias, with 15 studies missing on the left side of the funnel plot. After adjusting the SMD value for these studies, the effect size became −0.478.

**Figure 4:**
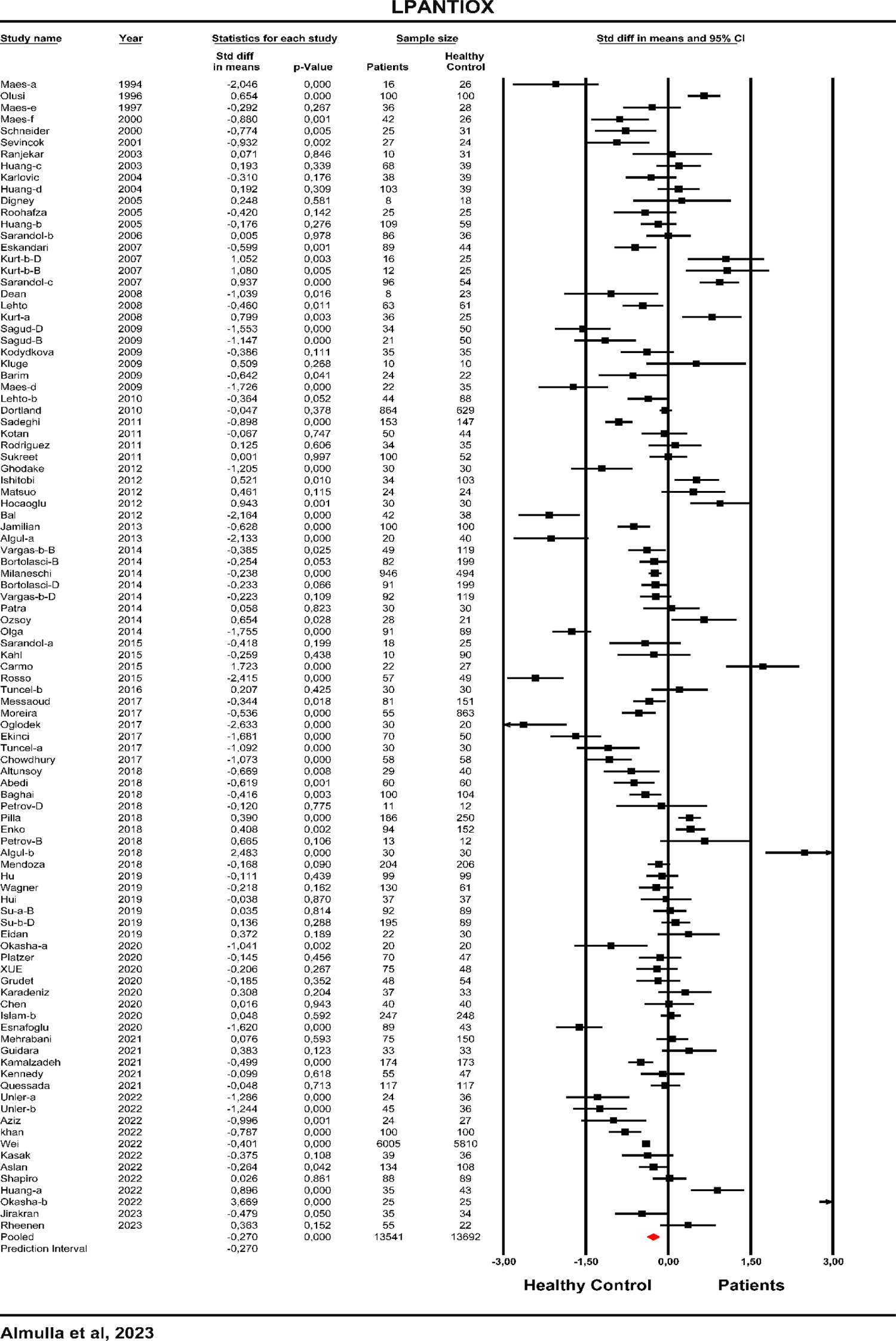
The forest plot of lipid-associated antioxidants (LPANTIOX) in patients with major depression and bipolar disorder as compared to healthy controls.

### LPOSTOX profile

In the present meta-analysis, we included 93 LPOSTOX studies. **Table 1** indicates that only 4 studies showed complete CIs on the negative side of zero, while 55 studies exhibited entire CIs on the positive side of zero. Additionally, zero-overlapping CIs were observed in 32 studies, and the SMD values were positive in 21 and negative in 11 studies. The forest plot displayed in **Figure 5** demonstrates a significantly increased LPOSTOX with large effect size (0.731) both in MDD and BD patients as compared with healthy controls. Table 3 reveals significant bias, as 18 studies were missing on the right side of the funnel plot. These missing studies were imputed, resulting in an even larger estimated effect size (0.987).

**Figure 5:**
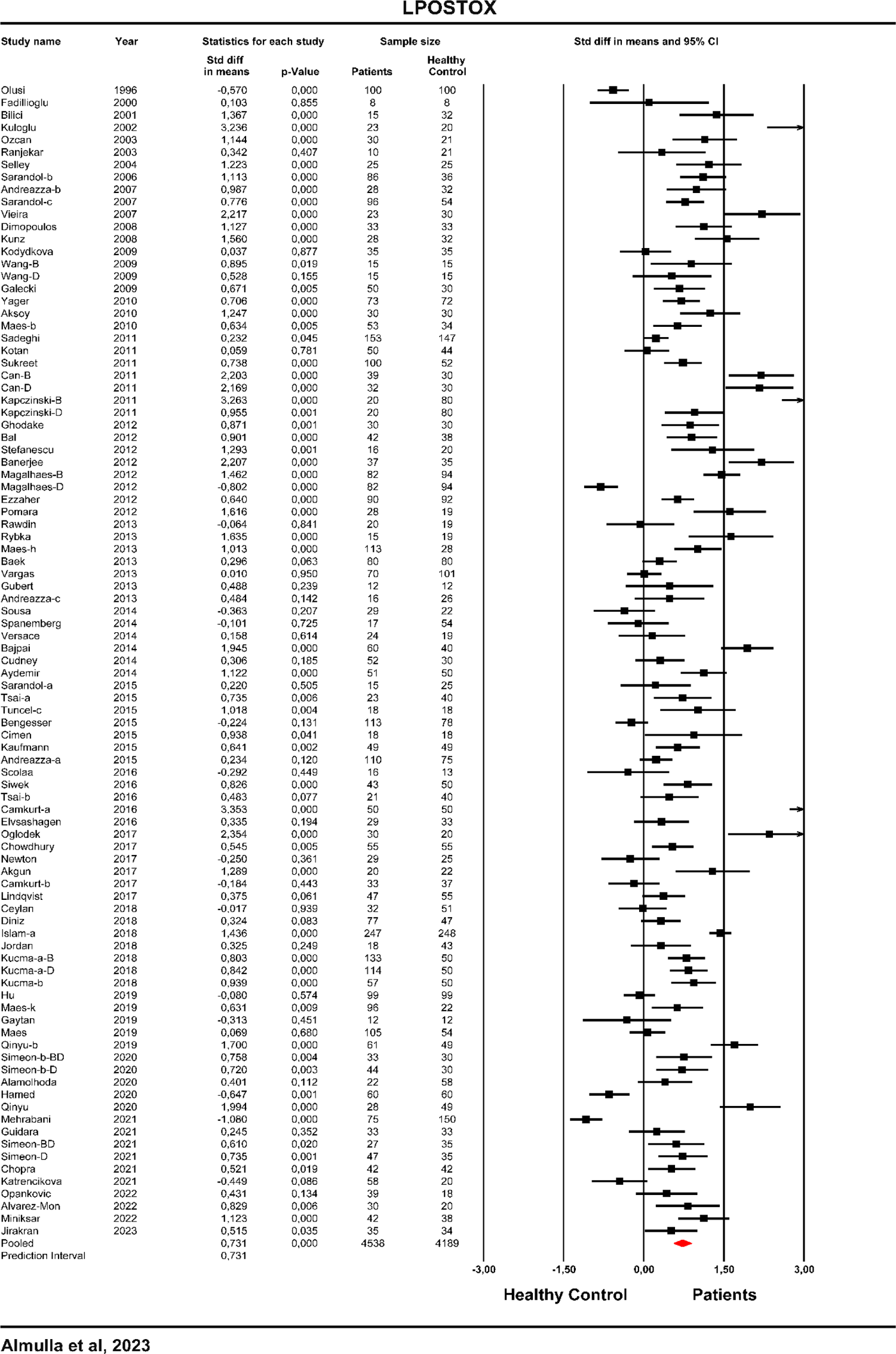
The forest plot of lipid-associated oxidative stress toxicity (LPANTIOX) in patients with major depression and bipolar disorder as compared to healthy controls.

### LPAUTO profile

The LPAUTO composite score, reflecting autoimmune responses against neoepitopes, was examined by combining 7 studies. The entire CIs of all studies appeared on the positive side of zero as shown in Table 1. Table 2 and **Figure 6** indicate that MDD and BD patients exhibit a significantly elevated LPAUTO index with high effect size (0.904). Table 3 demonstrated the absence of any apparent bias in the results.

**Figure 6:**
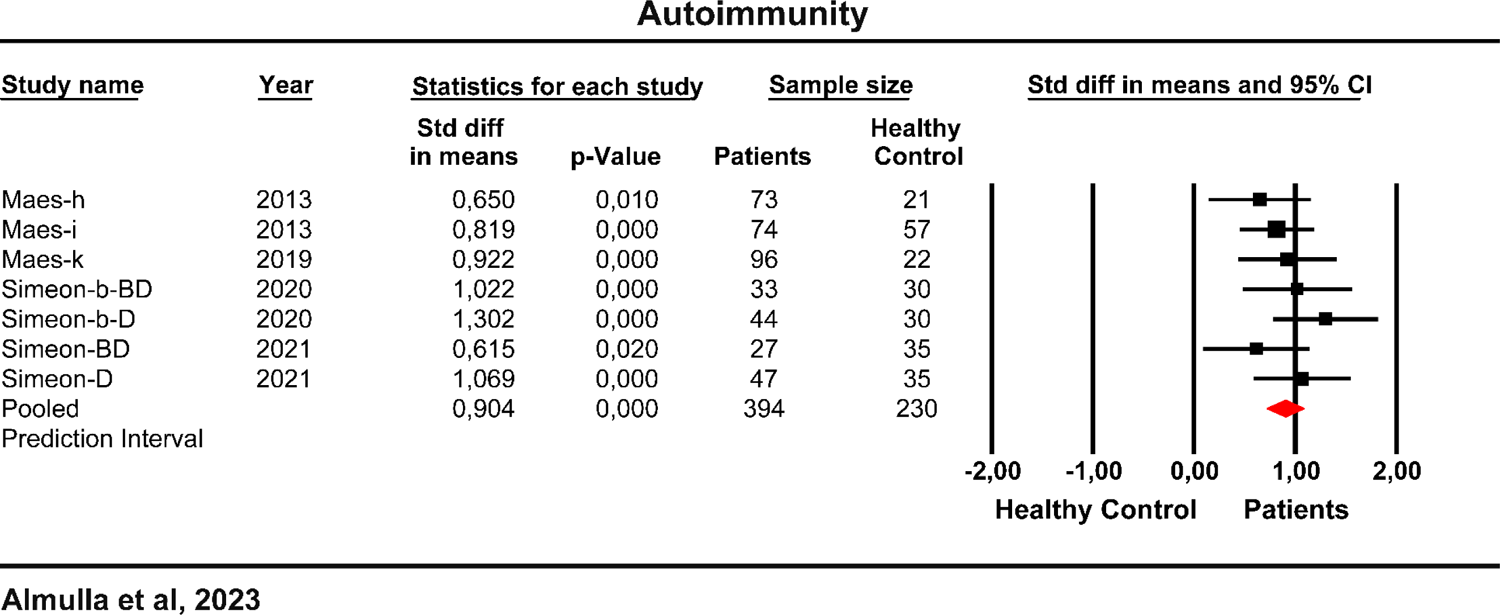
Forest plot of autoimmune responses to lipid-peroxidation neoepitopes in patients with major depression and bipolar disorder as compared to healthy controls.

### (LPOSTOX+LPAUTO) / LPANTIOX ratio

The present study has examined the (LPOSTOX+LPAUTO) / LPANTIOX ratio from 176 relevant studies. Of these studies, the entire CIs of 16 studies were negative, while it was completely positive in 89 studies. Furthermore, 71 studies exhibit a zero-intersection, and 27 studies had SMD values less than zero, while 44 studies had greater than zero SMD values. The group analysis showed a significant difference (p=0.027) in the (LPOSTOX+LPAUTO) / LPANTIOX ratio between patients with MDD and BD. However, Table 2 indicated that both MDD and BD patients demonstrated a significant increase in the former ratio, yet the effect size was greater in BD patients than in MDD patients. Table 3 revealed that in BD, there were 8 missing studies on the right side of the funnel plot and imputing these studies increases the estimated effects size (0.821). On the other hand, in MDD, there were 24 missing studies on the left side of the funnel plot, and adjusting the effect size to include these studies would reduce it to 0.141 (with 95% CI of 0.007; 0.274).

ESF, Tables 6-8 show the outcomes of the meta-analyses performed on the single indicators of the different indices used in the current study. ESF, Figures 1-8 show the forest plots of 8 solitary indicators. ESF, Listing show the list of all eligible studies.

### Meta-regression analyses

Meta-regression analyses were conducted to identify the single indicators that contribute to the heterogeneity observed in studies on lipid peroxidation and antioxidant defenses. The findings, as presented in ESF, Table 9, reveal that BD versus MDD exert a significant influence on several variables including ghrelin, 4-HNE, MDA, and the (LPOSTOX+LPAUTO) / LPANTIOX ratio. The latter, MDA and LPOSTOX were substantially affected by Young Mania Rating Scale (YMRS) scores. Moreover, latitude was identified as one of the most significant factors influencing RCT, HDL, LPANTIOX, and peroxides. On the other hand, body mass index significantly impacted vitamin E, MDA, and peroxides. Additionally, other factors such as age, gender, duration of illness, and age of onset demonstrate varying effects on different variables, all of which are presented in ESF, Table 9.

## Discussion

This is the first large-scale meta-analysis providing strong evidence for the role of RCT, lipid-associated antioxidant systems, lipid peroxidation, and autoimmunity to oxidative specific epitopes in the pathophysiology of MDD and BD. In fact, the current meta-analysis is the first to examine autoimmunity against oxidatively modified neoepitopes and all relevant RCT indicators.

### Reduced RCT in MDD and BD patients

The first major finding of the current meta-analysis is that MDD and BD are characterized by significantly reduced RCT as determined by HDL, PON1, Apo A and LCAT activity. The RCT pathway is a key antioxidant pathway that protects against atherogenicity, lipid peroxidation, inflammation and elevated bacterial load (Jirakran et al., 2023). Our results are in agreement with a recent pre-publication showing decreased RCT in MDD and suicidal behaviors (Jirakran et al., 2023). Maes et al. published the first study showing that MDD is accompanied by lowered RCT and elevated atherogenicity, as indicated by a reduced HDL/total cholesterol ratio (Maes et al., 1994). Further research also detected increased levels of atherogenic biomarkers in MDD patients (Bortolasci et al., 2015; Morelli et al., 2021; Nunes et al., 2015). Elevated atherogenicity coupled with oxidative and inflammatory mechanisms may explain, at least in part, the cooccurrence of MDD, atherosclerosis, and cardiovascular diseases (Maes et al., 2011e).

### Reduced vitamins A and D and coenzyme Q10 in MDD and BD

The second major finding of the current meta-analysis is that patients with MDD and BD have significantly decreased levels of lipid-soluble vitamins and coenzyme Q10. The analysis of the solitary components of this composite indicate a significant decrease of vitamin A and D, but not E, while there are no studies concerning vitamin K and only one study on CoQ10. Previous meta-analyses also detected lowered levels of some selected lipid-soluble vitamins. In this regard, Anglin et al. reported decreased vitamin D levels in MDD (Anglin et al., 2013), and Liu et al. found no significant changes in vitamin E (Liu et al., 2015). Nevertheless, not one of the prior meta-analysis has covered all lipid-soluble vitamins in MDD and BD patients. Thus, the current study provides the first proof that not only RCT-related antioxidant defenses but also the lipid-soluble vitamins and coenzyme Q10 are lowered in affective disorders.

So, one of the most important things we learned from our study is that both MDD and BD are linked to a weakening of the antioxidant defense system associated with lipids, and that there are no major differences between MDD and BD in terms of lipid antioxidants except for the levels of ghrelin.

#### Increased lipid peroxidation in MDD and BD

The third major finding of the present study is that MDD and BD are characterized by an overall increase in oxidative damage to lipids, as reflected by the elevated LPOSTOX index. MDA, 4-HNE, and isoprostanes were the most essential lipid-associated peroxidation biomarkers. Intriguingly, the effect size of MDA was significantly greater in BD than in MDD. Nevertheless, there were no significant changes in LOOH, Apo B, or oxLDL in affective disorders, whilst other biomarkers, such as 7-ketocholesterol and hydroxycholesterol, could not be individually examined since they were assessed in one study only.

Our LPOSTOX results extend those of previous studies (Black et al., 2015; Brown et al., 2014; Jiménez-Fernández et al., 2021, 2022; Liu et al., 2015; Mazereeuw et al., 2015). Nevertheless, the analysis of Liu, Zhong et al. (2015) was based mainly on a Chinese population, that may have restricted the generalizability of the results since there are ethnical differences in oxidative stress responses (Morris, Zhao et al. 2012). Moreover, all previous meta-analyses were performed on solitary biomarkers rather than examining the more important overall effect of lipid peroxidation as assessed by combining all relevant biomarkers.

#### Increased autoimmune responses to oxidative specific epitopes

The fourth major finding of the present study is that affective disorders are accompanied by increased autoimmune responses against neoepitopes generated by oxidative stress. These findings indicate increased oxidative stress, lipid peroxidation, damage to LDL, aldehyde formation, and an autoimmune response directed to the oxidatively modified epitopes namely oxLDL and MDA (Maes et al., 2013a; Maes et al., 2013b; Simeonova et al., 2021b; Simeonova et al., 2020a). The latter neoepitopes have proinflammatory effects, and are additionally immunogenic, thereby mounting autoimmune responses (Maes et al., 2013b). Moreover, these oxidative-specific epitopes in MDD and BD may act as damage-associated molecular patterns, which activate the Toll-Like Receptor (TLR)–Radical Cycle, thereby provoking and maintaining immune-inflammatory responses (Lucas and Maes, 2013).

All in all, affective disorders are accompanied by increased lipid peroxidation and autoimmune response and decreased lipid antioxidant defenses, as indicated by the (LPOSTOX + LPAUTO) / LPANTIOX ratio. Interestingly, the degrees of lipid peroxidation may be somewhat higher in BD than MDD.

#### Consequences for the immune-inflammatory theory of MDD/BD

A decreased RCT is a key factor in MDD and BD because this condition increases the vulnerability to developing lipid peroxidation, oxidative damage of lipids, and, therefore, autoimmunity to oxidative specific epitopes, inflammation and increased atherogenicity (Marchio et al., 2019; Morris et al., 2021b). The PON1 Q192R polymorphism determines in part PON1 activity and, therefore, plays a key role in oxidative and inflammatory stress in MDD/BD (Bortolasci et al., 2014). HDL, PON1 and ApoA tightly regulate immune responses (Morris et al., 2021a; Morris et al., 2020a; Morris et al., 2019; Morris et al., 2020b; Morris et al., 2021c): a) HDL inhibits NF-κB activity, promotes dendritic cell maturation and antigen presentation to T cells, b) lowers TLR-4 activation by removing cholesterol from membrane lipid rafts; c) HDL affects the complement system, chemotaxis of monocytes and macrophages, and the activation of T cells and B cells; and d) HDL has a key role in reducing inflammation and suppressing the immune system by degrading membrane lipid rafts and activating pentraxin 3, a molecule that plays a fundamental role in the immune response (Morris et al., 2022). PON1 protects against oxidative damage to immune cell membranes, oxidized lipoproteins, and mitochondria, upregulates glucose transporter-1 (GLUT-1), and regulates aerobic glycolysis, the pentose monophosphate pathway, fatty acid oxidation, and peroxisome proliferator-activated receptor gamma (PPAR-γ) activity (Morris et al., 2021a; Morris et al., 2020a). ApoA1 regulates the Th-17/Treg balance, enhances mitochondrial functions, activates the electron transport chain and stabilizes PON1 in HDL to preserve its activity (Morris et al., 2022).

Vitamins A and E serve as antioxidants in cell membranes and lipoproteins by inhibiting ROS and neutralizing free radicals (Burton et al., 1983; Palace et al., 1999; Popa et al., 2021). While vitamin D may not scavenge free radicals like vitamins E and C, it upregulates the master antioxidant gene, Nrf2, and antioxidant enzymes such as glutathione peroxidase, which both protect against oxidative damage (Boaventura and Cembranel, 2020; Wimalawansa, 2019). In addition, vitamin D promotes an energy-conserving state within mitochondria by stimulating processes such as fusion, mitophagy, and biogenesis, which in turn may renew mitochondria, maintain energy metabolism, and reduce oxidative stress (Bhutia, 2022; Sheeley et al., 2022). Coenzyme Q10 is known for its potent antioxidant, anti-inflammatory and neuroprotective properties (Maes et al., 2009; Morris et al., 2013). Moreover, Coenzyme Q10 lowers NF-κB-gene expression and the production of pro-inflammatory cytokines, such as tumor necrosis factor (TNF)-α, and protects against lipopolysaccharide (LPS)-induced inflammatory responses (Abd El-Gawad and Khalifa, 2001; Schmelzer et al., 2008; Schmelzer et al., 2007a; Schmelzer et al., 2007b; Sugino et al., 1987).

Oxidative stress, including lipid peroxidation, augments inflammatory processes by activating redox-sensitive transcription factors, like NF-κB, causing expression of pro-inflammatory genes and the release of inflammatory signaling molecules (Kabe et al., 2005; Morris et al., 2022). In this respect, some studies established activated NF-κB in MDD and BD (Miklowitz et al., 2016; Roman et al., 2021). Furthermore, the nucleotide-binding domain, leucine-rich–containing family, pyrin domain–containing-3 (NLRP3) inflammasome, a key molecule in initiating innate immunity, is also activated by oxidative stress pathways (Morris and Maes, 2014; Zhou et al., 2011). Increased lipid peroxidation also causes depletion of ω3-polyunsaturated fatty acids (PUFA) in plasma cholesteryl esters and phospholipids, which in turn increases the production of pro-inflammatory cytokines, especially in response to psychological stressors (Maes et al., 2000a; Maes et al., 1999).

Increased oxidative stress may cause damage to enzymes of the electron transport chain, structural and functional phospholipids, including cardiolipin (Kakkar and Singh, 2007; Morris and Maes, 2014; Paradies et al., 2011; Pope et al., 2008; Turrens, 2003). Furthermore, lipid peroxidation products, such as MDA, are neurotoxic by altering mitochondrial proteins resulting in mitochondrial dysfunction within the neurons and, hence, and inducing impairments in energy production (Long et al., 2009), which have been reported in mood disorders (Caruso et al., 2019). The resulting aberrations in ATP generation may further accelerate ROS production, thereby establishing a self-sustaining cycle of increasing neurotoxicity (Kakkar and Singh, 2007; Morris and Maes, 2014; Turrens, 2003). Due to a high need for oxygen and high levels of PUFAs, brain cells are more likely to be damaged by the neurotoxic effects of lipid peroxidation. This makes neuronal cells highly vulnerable to oxidative stress-induced neurotoxicity and neurodegeneration (Syafrita et al., 2020).

Our results suggest that novel treatments of affective disorders should target the intertwined aberrations in oxidative and immune-inflammatory pathways. Some available antidepressants show a potential to lower oxidative stress (Maes et al., 2012). For example, Ahmadimanesh et al. found that selective serotonin reuptake inhibitors (SSRIs) significantly decreased DNA damage by reducing 8-hydroxy-2-deoxyguanosine (8-OHdG) and IL-6 (Ahmadimanesh et al., 2019). Furthermore, Kotan et al., in a longitudinal study, found that antidepressant treatments significantly decrease plasma MDA and increase PON1 activities (Kotan et al., 2011).

### Limitation

When evaluating the results of this research, it is important to take into account a number of limitations. Higher heterogeneity caused by some identified input variables may have affected the precision and accuracy of the pooled effect estimates. Latitude was one of the primary drivers of this increased variability, and we identified a substantial negative relationship between latitude and RCT, HDL, and LPANTIOX, and a positive relationship with peroxides. Second, the limited studies regarding lipid peroxidation biomarkers in the brain and CSF restrict our ability to complete the profile of central lipid peroxidation. However, our group analysis showed no significant difference between central and peripheral LPOSTOX composite scores. Therefore, more post-mortem and CSF studies are needed to examine the status of central lipid peroxidation and antioxidant biomarkers. Third, the absence of data on vitamin K in MDD and BD, and the paucity of studies on Coenzyme Q10 limit our ability to construct a thorough lipid-soluble vitamin (ADEKQ) profile and conduct separate meta-analyses. Fourth, the limited information regarding metabolic syndrome, which impacts HDL, ApoA, PON1 and some lipid peroxidation biomarkers, prevented us to examine the effects of metabolic syndrome (Jirakran et al., 2023). Fifth, given that most studies did not disclose data about antidepressant and mood stabilizer usage, we were unable to investigate their impact on lipid peroxidation biomarkers. Thus, more studies are required on drug-naïve versus treated MDD and BD patients.

## Conclusions

The present large-scale meta-analysis shows that patients with MDD and BD have a decreased RCT and other lipid-associated antioxidant defenses coupled with higher oxidative damage to lipids, and greater IgG and IgM-mediated autoimmune responses to oxidatively modified neoepitopes. **Figure 7** summarizes the importance of increased lipid peroxidation and decreased lipid-associated antioxidant defenses for the enhanced inflammatory and autoimmune responses in MDD/BD. Hence, targeting these pathways is crucial for the treatment of MDD and BD.

**Figure 7:**
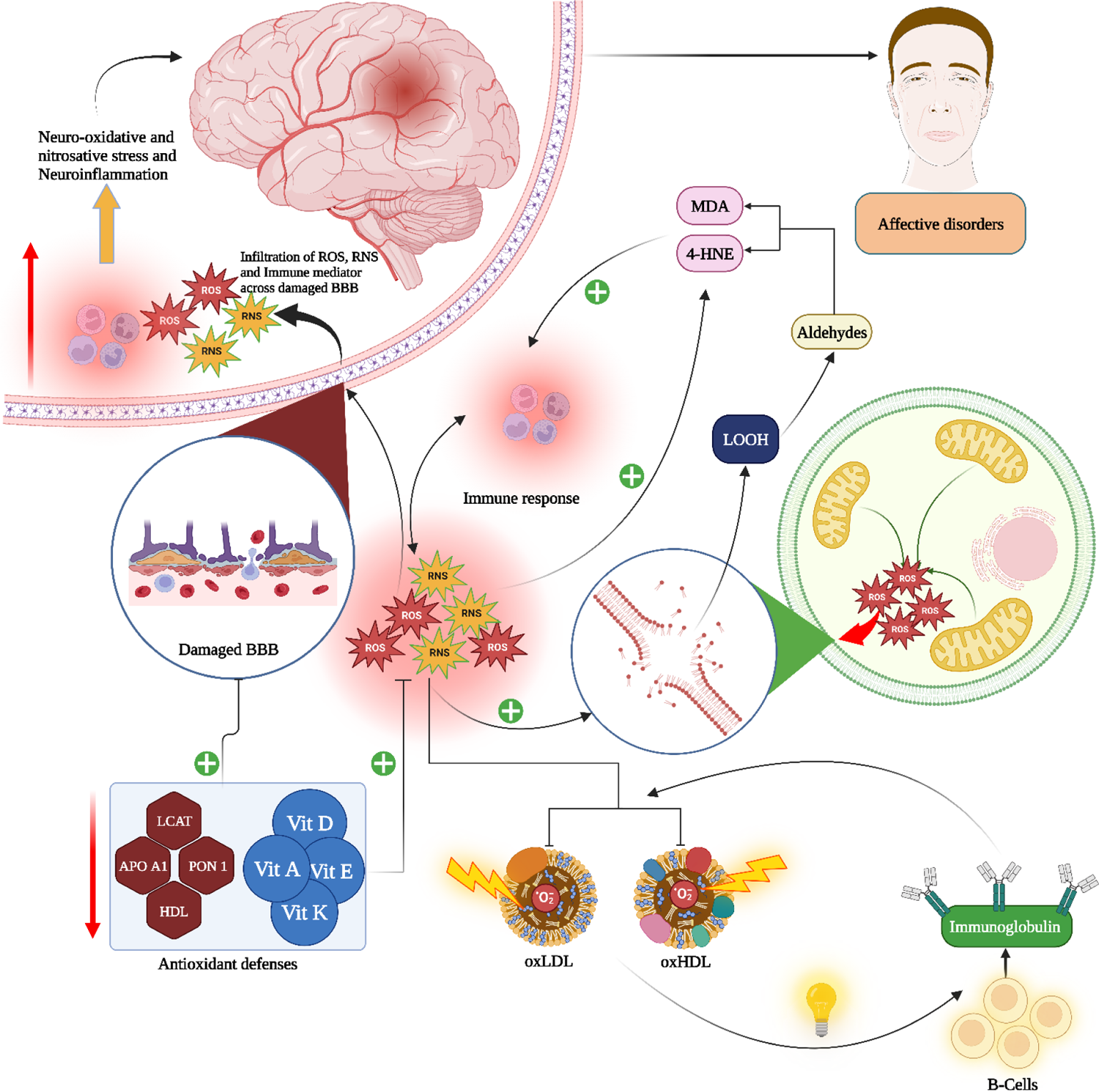
Summary of the role of reverse cholesterol transport (RCT), decreased lipid antioxidants and increased lipid peroxidation in the pathophysiology of major depression and bipolar disorder

## Declaration of Competing Interests

The authors have no conflicts of interest to declare.

## Ethical approval and consent to participate

Not applicable.

## Consent for publication

Not applicable.

## Availability of data and materials

The last author (MM) will respond to any reasonable requests for the dataset utilized in the present meta-analysis, following its full exploitation by all authors. This dataset will be provided in Excel file format.

## Funding

The study was funded by the C2F program, Chulalongkorn University, Thailand, No. 64.310/436/2565 to AFA, and an FF66 grant and a Sompoch Endowment Fund (Faculty of Medicine), MDCU (RA66/016) to MM.

## Author’s contributions

The design of the current study is performed by AA and MM. AA and YT collected the data. AA and MM conducted the statistical analysis. All authors participated in the writing and revision of the manuscript and have approved the submission of the final draft.

## Acknowledgments

Not applicable.

## Notes

### Competing Interest Statement

The authors have declared no competing interest.

