## supplementary file for "Reverse cholesterol transport and lipid peroxidation biomarkers in major depression and bipolar disorder: a systematic review and meta-analysis"

SHORT TITLE: Lipid peroxidation in affective disorders

Abbas F. Almulla, Ph.D.[a,b] , Yanin Thipakorn, M.D., Ph.D.[a] , Ali Abbas Abo Algon, M.Sc. [c], Chavit Tunvirachaisakul, M.D., Ph.D.[a,d] , Hussein K. Al-Hakeim, Ph.D.[e], Michael Maes, M.D., Ph.D.[a,f,g,h]

[a] Department of Psychiatry, Faculty of Medicine, Chulalongkorn University, Bangkok, Thailand.

[b] Medical Laboratory Technology Department, College of Medical Technology, The Islamic University, Najaf, Iraq.

[c] Iraqi Education Ministry, Najaf, Iraq

[d] Cognitive Impairment and Dementia Research Unit, Faculty of Medicine, Chulalongkorn University, Bangkok, Thailand.

[e] Department of Chemistry, College of Science, University of Kufa, Kufa, Iraq.

[f] Department of Psychiatry, Medical University of Plovdiv, Plovdiv, Bulgaria.

[g] Department of Psychiatry, IMPACT Strategic Research Centre, Deakin University, Geelong, Victoria, Australia.

[h] Kyung Hee University, 26 Kyungheedae-ro, Dongdaemun-gu, Seoul 02447, Korea

**Corresponding author:**

Prof. Dr Michael Maes, M.D., Ph.D.

Department of Psychiatry

Faculty of Medicine, Chulalongkorn University

Bangkok, 10330

Thailand

E-mail addresses:

#### ESF, Table 1. Search sentences and terms used in each database

| Database Name | Search Sentence | No. of Articles |
|---|---|---|
| PubMed/Medline | ((((((((((((Depression and Lipid peroxidation)* OR (Major depression and Malonaldehyde)) OR (Bipolar disorder and Lipid peroxidation)) OR (Depression and Isoprostane)) OR (Bipolar disorder and Isoprostane)) OR (Bipolar disorder and Malonaldehyde)) OR (Depression and oxidizedLDL)) OR (Bipolar disorder and oxidizedLDL)) OR (Depression and TBARS)) OR (Bipolar and TBARS)) OR (Depression and 4-HNE)) OR (Bipolar and 4-HNE)) OR (MDD and Lipid peroxidation)) OR (Ketocholesterol and Depression) | **1838** |
| | ((((((((((MDA and Depression) OR (Malonaldehyde and MDD)) OR (IsoPs and Depression)) OR (F2 isoprostanes and Depression)) OR (F2 isoprostanes and MDD)) OR (Bipolar disorder and F2 isoprostanes)) OR (Bipolar disorder and MDA)) OR (Bipolar disorder and Lipid peroxidation)) OR (MDD and Lipid peroxidation)). | **1363** |
| | (((((((((HDL and Depression) OR (HDL and Bipolar disorder)) OR (Lipid antioxidants and Depression)) OR (Depression and Paroxenase 1)) OR (Bipolar disorder and Paroxenase 1)) OR (Depression and LCAT)) OR (Depression and Apo A)) OR (Depression and Apo E)) OR (Depression and Apo D)) OR (Depression and Apo B). | **3650** |
| | ((((Depression and LCAT)) OR (Depression and Apo A)) OR (Depression and Apo E)) OR (Depression and Apo D)) OR (Depression and Apo B). | **682** |
| | (((((Depression and Vitamin D)) OR (Depression and Vitamin E)) OR (Depression and Vitamin A)) OR (Depression and Vitamin K)) OR (Depression and Ghrelin) | **3149** |
| | (((((Bipolar disorder and Vitamin D)) OR (Bipolar disorder and Vitamin E)) OR (Bipolar disorder and Vitamin A)) OR (Bipolar disorder and Vitamin K)) OR (Bipolar disorder and Ghrelin) | **111** |

| | | |
|---|---|---|
| **Google Scholar** | ((((((((Depression* and Lipid peroxidation) OR (MDA)) OR (Isoprostane)) OR (4-HNE)) OR (LOOH)) OR (Peroxides)) OR (oxidizedLDL)) OR (oxidizedHDL)) OR (TBARS)) AND ((((((((Bipolar disorder * and Lipid peroxidation) OR (MDA)) OR (Isoprostane)) OR (4-HNE)) OR (LOOH)) OR (Peroxides)) OR (oxidizedLDL)) OR (oxidizedHDL)) OR (TBARS)) | **13000** |
| | ((((((((Depression* and Lipid Antioxidants) OR (PON1)) OR (LCAT)) OR (CoQ10)) OR (HDL) OR (vitamin A)) OR (vitamin E)) OR (vitamin K)) OR (Ghrelin)) OR (Apo A)) OR (Apo E)) OR (Apo D)) OR (Apo B)) AND ((((((((Bipolar disorder * and Lipid Antioxidants) OR (PON1)) OR (LCAT)) OR (CoQ10)) OR (HDL)) OR (vitamin A)) OR (vitamin E)) OR (vitamin K)) OR (Ghrelin)) OR (Apo A)) OR (Apo E)) OR (Apo D)) OR (Apo B)) | **18300** |
| **SciFinder** | Depression OR Bipolar disorder and Malonaldehyde and TBARS, Bipolar disorder and TBARS, and 4-HNE, and LOOH (20) and LOOH , oxLDL, and oxLDL. | **3233** |
| | Depression OR Bipolar disorder and PON1 and LCAT and HDL and Apo A and Apo E and Apo B and CoQ10, vitamin D and vitamin E and vitamin K and vitamin A. | 10828 |

**ESF, Table 2. Immune cofounder's scale (ICS) applied from Andrés-Rodríguez, et al., 2019**

| **Methodological quality of the study** | |
|---|---|
| **1** | Study sample ≥ 128 participants including patients and controls (1= Yes, 0 = No) |
| **2** | Did the study control the results for potential confounders (e.g., age, BMI, gender, race)? (1= Yes, 0 = No) |
| **3** | Were participants with schizophrenia and controls age- and-gender-matched or was there a statistical control? (1= Yes, 0 = No) |
| **4** | Was the time of sample collection specified (e.g., morning vs. evening)? (1= Yes, 0 = No) |
| **5** | Were participants with Alzheimer disease free of immunomodulatory drugs including anti-cytokines, glucocorticoids, immunoglobulins, and immunosuppressants, or was there a medication washout period or was drug intake statistically controlled for? (1= Yes, 0 = No) |
| **6** | Were participants with schizophrenia free of nervous system drugs or were the data statistically controlled for? (1= Yes, 0 = No) |
| **7** | Reporting either the manufacturer of the test or detection limit and coefficients of variation (1= Yes, 0 = No) |
| **8** | Reporting how data under detection limit were handled (1 = Yes, 0 = No) |
| **9** | Reporting % of the sample under detection limit (1=Yes, 0= No) |
| **10** | Reporting blood fraction (serum, plasma, culture supernatant or whole blood) (1= Yes, 0 = No) |
| | **Total quality score (10 points)** |
| **Biomarker confounders red points** | |

| *The red points should not be given if the item is statistically controlled for* | |
|---|---|
| **1** | 3 red points for comorbid illnesses such as autoimmune disorders & other immune disorders including rheumatoid arthritis, psoriasis, inflammatory bowel disease, chronic obstructive pulmonary disease, multiple sclerosis |
| **2** | 3 red points for use of recreational drugs such as methamphetamine or opioids |
| **3** | 2 red points when groups were not matched for age |
| **4** | 2 red points when groups were not matched for sex |
| **5** | 2 red points for medication use as for example immunomodulators |
| **6** | 2 red points for early traumatic life events |
| **7** | 2 red points for shift work and primary sleep disorders |
| **8** | 1.5 red points for use of neuroleptics |
| **9** | 1 red point for more common systemic immune disorders including diabetes type 1/2, essential hypertension, metabolic syndrome |
| **10** | 1 red point for not fasting (8 hours before blood extraction) |
| **11** | 1 red point for use of omega-3 and antioxidant supplements |
| **12** | 1 red point when data were not controlled for body mass index |
| **13** | 1 red point when data were not controlled for physical activity or sedentary life |
| **14** | 1 red point when data were not controlled for smoking |

| | |
|---|---|
| **15** | 1 red point for use of oral contraceptives or NSAIDs |
| **16** | 0.5 red points when data were not controlled for ethnicity in countries such as US, Brazil |
| **17** | 0.5 red points when data were not controlled for seasonality |
| **18** | 0.5 red points when data were not controlled for diurnal variation (8-10 a.m. versus all other time points) |
| | **Total red point score (26 points)** |

**ESF, Table 3. PRISMA checklist**

| Section/topic | # | Checklist item | Reported on page # |
|---|---|---|---|
| **TITLE** | | | |
| Title | 1 | Identify the report as a systematic review, meta-analysis, or both. | 1 |
| **ABSTRACT** | | | |
| Structured summary | 2 | Provide a structured summary including, as applicable: background; objectives; data sources; study eligibility criteria, participants, and interventions; study appraisal and synthesis methods; results; limitations; conclusions and implications of key findings; systematic review registration number. | 3 |
| **INTRODUCTION** | | | |
| Rationale | 3 | Describe the rationale for the review in the context of what is already known. | 5-8 |
| Objectives | 4 | Provide an explicit statement of questions being addressed with reference to participants, interventions, comparisons, outcomes, and study design (PICOS). | 8 |
| **METHODS** | | | |
| Protocol and registration | 5 | Indicate if a review protocol exists, if and where it can be accessed (e.g., Web address), and, if available, provide registration information including registration number. | 8 |
| Eligibility criteria | 6 | Specify study characteristics (e.g., PICOS, length of follow-up) and report characteristics (e.g., years considered, language, publication status) used as criteria for eligibility, giving rationale. | 9 |
| Information sources | 7 | Describe all information sources (e.g., databases with dates of coverage, contact with study authors to identify additional studies) in the search and date last searched. | 9 |
| Search | 8 | Present full electronic search strategy for at least one database, including any limits used, such that it could be repeated. | 9 |
| Study selection | 9 | State the process for selecting studies (i.e., screening, eligibility, included in systematic review, and, if applicable, included in the meta-analysis). | 9,10 |

| | | | |
|---|---|---|---|
| Data collection process | 10 | Describe method of data extraction from reports (e.g., piloted forms, independently, in duplicate) and any processes for obtaining and confirming data from investigators. | 10,11 |
| Data items | 11 | List and define all variables for which data were sought (e.g., PICOS, funding sources) and any assumptions and simplifications made. | ESF, Table 1 |
| Risk of bias in individual studies | 12 | Describe methods used for assessing risk of bias of individual studies (including specification of whether this was done at the study or outcome level), and how this information is to be used in any data synthesis. | 12,13 and Table 3 |
| Summary measures | 13 | State the principal summary measures (e.g., risk ratio, difference in means). | 12,13 |
| Synthesis of results | 14 | Describe the methods of handling data and combining results of studies, if done, including measures of consistency (e.g., $I^2$) for each meta-analysis. | 12,13 |
| Risk of bias across studies | 15 | Specify any assessment of risk of bias that may affect the cumulative evidence (e.g., publication bias, selective reporting within studies). | 13 and Table 3 |
| Additional analyses | 16 | Describe methods of additional analyses (e.g., sensitivity or subgroup analyses, meta-regression), if done, indicating which were pre-specified. | 13, 16 and ESF, Table 9 |
| **RESULTS** | | | |
| Study selection | 17 | Give numbers of studies screened, assessed for eligibility, and included in the review, with reasons for exclusions at each stage, ideally with a flow diagram. | 13, 14 |
| Study characteristics | 18 | For each study, present characteristics for which data were extracted (e.g., study size, PICOS, follow-up period) and provide the citations. | ESF, Table 5 |
| Risk of bias within studies | 19 | Present data on risk of bias of each study and, if available, any outcome level assessment (see item 12). | Table 3 |
| Results of individual studies | 20 | For all outcomes considered (benefits or harms), present, for each study: (a) simple summary data for each intervention group (b) effect estimates and confidence intervals, ideally with a forest plot. | Table 1 |
| Synthesis of results | 21 | Present results of each meta-analysis done, including confidence intervals and measures of consistency. | Table 2 |
| Risk of bias across studies | 22 | Present results of any assessment of risk of bias across studies (see Item 15). | Table 3 |
| Additional analysis | 23 | Give results of additional analyses, if done (e.g., sensitivity or subgroup analyses, meta-regression [see Item 16]). | 17 |

| | | | |
|---|---|---|---|
| **DISCUSSION** | | | |
| Summary of evidence | 24 | Summarize the main findings including the strength of evidence for each main outcome; consider their relevance to key groups (e.g., healthcare providers, users, and policy makers). | 18-24 |
| Limitations | 25 | Discuss limitations at study and outcome level (e.g., risk of bias), and at review-level (e.g., incomplete retrieval of identified research, reporting bias). | 24 |
| Conclusions | 26 | Provide a general interpretation of the results in the context of other evidence, and implications for future research. | 25 |
| **FUNDING** | | | |
| Funding | 27 | Describe sources of funding for the systematic review and other support (e.g., supply of data), role of funders for the systematic review. | 26 |

**EFS, Table 4. Studies excluded from the meta-analysis but included in the systematic review**.

| **Authors, year** | **Reason why excluded from the meta-analysis** |
|---|---|
| Kalenderolu et al. 2010 | No data of the measured biomarker of healthy control |
| Cusa et al. 2013 | Outlier |
| Khanzode et al. 2013 | Outlier |
| Kilicarslan et al. 2022 | Outlier |
| Mondin et al. 2016 | Outlier |
| Chung et al. 2013 | Conducted on urine sample |

#### ESF, Table 5. Characteristics of studies which were included in the systematic reviews and meta-analysis

| # | Authors, years | Setting | Type of case | Type of Control | Sample Size | | | Age | | Assessed Biomarkers | Specimen | Method | Quality score | Red point score | Findnigs in MDD and BD patients compared to HC |
|---|---|---|---|---|---|---|---|---|---|---|---|---|---|---|---|
| | | | | | Cases (M/F) | Control M/F | Total M/F | Case-Mean (SD) | Control-Mean(SD) | | | | | | |
| 1 | (Van Rheenen, Ringin et al. 2023) | Australia | BD | HC | 55 (32/23) | 22 (13/9) | 77 (45/32) | 37.8 (11.3) | 35.2(10.9) | Vitamin D | Plasma | ELISA | 5 | 10 | Vitamin D # |
| 2 | (Yıldız Miniksar and Göçmen 2022) | Turkey | MDD | HC | 42 (12/30) | 38 (14/24) | 80 (26/54) | 15.48 (1.58) | 14.42 (2.99) | MDA | Serum | Spectro. | 6 | 9,5 | MDA # |
| 3 | (Kilicarslan, Sahan et al. 2022) | Turkey | BD | HC | 50 (14/36) | 50 (16/34) | 100 (30/70) | 39.12 (10.19) | 38.18 (11.02) | 4-HNE | Serum | Spectro. | 5 | 12 | 4-HNE# |
| 4 | (Wei, Wang et al. 2022) | China | BD | HC | 6005 (3178/2827) | 5810 (2850/2960) | 11815 (6028/5787) | 38.89 (0.168) | 38.54 (0.145) | HDL | Blood | Unknown | 6 | 9,5 | HDL* |
| 5 | (Ünler, Ekmekçi Ertek et al. 2022) | Turkey | BD | HC | 24 (9/15) | 36 (7/29) | 60 (16/44) | 41.08 (12.92) | 40.83 (10.21) | Ghrelin | Serum | ELISA | 3 | 11 | Ghrelin* |
| | | | UD | | 45 (8/37) | | 81 (15/66) | 44.22 (10.44) | 40.83 (10.21) | | | | | | Ghrelin* |
| 6 | (Shapiro, Kennedy et al. 2022) | Canada | BD | HC | 88 (31/57) | 89 (50/39) | 177 (81/96) | 17.5 (1.7) | 17.2 (1.8) | HDL | Blood | Automated | 6 | 7 | HDL# |
| 7 | (Opanković, Milovanović et al. 2022) | Serbia | MDD | HC | 39 (7/32) | 18 (5/13) | 57 (12/45) | 49.5 (11.8) | 42.5 (7.7) | MDA | Serum | ELISA | 4 | 10 | MDA # |
| 8 | (Okasha, El-Gabry et al. 2022) | Egypt | MDD | HC | 25 (17/8) | 25 (17/8) | 50 (34/16) | 34.56 (11.47) | 0 (0) | Ghrelin | Serum | ELISA | 5 | 8 | Ghrelin # |
| 9 | (khan, Shafiq et al. 2022) | Pakistan | Depression | HC | - | - | 0 (0/0) | - | - | Vitamin D | Serum | Automated | 5 | 11,5 | Vitamin D * |
| 10 | (Kasak, Ceylan et al. 2022) | Turkey | BD | HC | 39 (12/27) | 36 (9/27) | 75 (21/54) | 16.7 (1.27) | 17.0 (0.75) | HDL | Serum | Unknown | 6 | 7,5 | HDL * |
| 11 | (Huang, Chen et al. 2022) | Taiwan | BD | HC | 38 (19/19) | 43 (16/27) | 81 (35/46) | 28.63 (3.82) | 26.16 (4.04) | Ghrelin | Serum | Immunoassay | 5 | 9,5 | Ghrelin # |
| | | | | | 31 (7/24) | | 74 (23/51) | 25.26 (5.39) | 26.16 (4.04) | | | | | | Ghrelin # |
| 12 | (Draghici 2022) | Romania | Mood disorder | HC | 134 (19/115) | 108 (14/94) | 242 (33/209) | 65.67 (6.56) | 65.25 (7.17) | HDL | Serum | Automated | 6 | 9 | HDL * |
| 13 | (Alvarez-Mon, Ortega et al. 2022) | Spain | MDD | HC | 30 (19/11) | 20 (12/8) | 50 (31/19) | 43.26 (13.14) | 40.45 (12.46) | MDA | Plasma | Spectro. | 6 | 7,5 | MDA # |

| | | | | | | | | | | | | | | | |
|---|---|---|---|---|---|---|---|---|---|---|---|---|---|---|---|
| 14 | (Abdel Aziz, Al-Mugaddam et al. 2022) | UAE | BD | HC | 24 (15/9) | 27 (13/14) | 51 (28/23) | 31.92 (9.789) | 34.41 (9.316) | Ghrelin | Serum | ELISA | 5 | 8 | Ghrelin * |
| 15 | (Vaghef-Mehrabani, Izadi et al. 2021) | Iran | MDD | HC | 75 (0/75) | 150 (0/150) | 225 (0/225) | 39.64 (7.70) | 39.97 (7.58) | HDL, MDA | Serum | Spectro. | 7 | 4,5 | HDL *, MDA # |
| 16 | (Quessada, Nascimento et al. 2021) | Brazil | Depression | HC | 117 (39/78) | 117 (48/69) | 234 (87/147) | 69.8 (8.1) | 71.5 (8.4) | Vitamin D | Plasma | Automated | 6 | 8,5 | Vitamin D * |
| 17 | (Platzer, Fellendorf et al. 2021) | Austria | BD | HC | 74 (74/0) | 35 (35/0) | 109 (109/0) | 46.5 (14.1) | 39.4 (16.1) | Ghrelin | Plasma | ELISA | 6 | 8 | Ghrelin # |
| | | | | | 65 (0/65) | 58 (0/58) | 123 (0/123) | 43.9 (13.7) | 38.3 (15.7) | | | | | | Ghrelin * |
| 18 | (Parul Chopra 2021) | India | MDD | HC | 42 (25/17) | 42 (23/19) | 84 (48/36) | 19.81 (4.26) | 20.62 (4.0) | 8-Isoprostane | Plasma | Immunoassay | 5 | 9 | 8-Isoprostane # |
| 19 | (Kennedy, Islam et al. 2021) | Canada | BD | HC | 55 (20/35) | 47 (23/24) | 102 (43/59) | 17.64 (1.76) | 17.34 (1.69) | HDL | Blood | Automated | 4 | 10,5 | HDL * |
| 20 | (Katrenčíková, Vaváková et al. 2021) | Slovakia | Depressive disorder | HC | 58 (12/46) | 20 (8/12) | 78 (20/58) | 15.6 (1.6) | 14.8 (2.4) | Peroxide | Serum | Spectro. | 6 | 6,5 | Peroxide * |
| 21 | (Kamalzadeh, Saghafi et al. 2021) | Iran | Depression | HC | 174 (14/160) | 173 (43/130) | 347 (57/290) | 42 (10) | 37 (10) | Vitamin D | Serum | Automated | 6 | 9,5 | Vitamin D * |
| 22 | (Guidara, Messedi et al. 2021) | Tunisia | BD | HC | 33 (33/0) | 40 (40/0) | 73 (73/0) | 32.00 (27.75–41.0) | 36.50 (31–40.5) | 24-OHCHO<br>25-OHCHO<br>27-OHCHO<br>7-ketocholesterol<br>HDL | Serum | UPLC-MS/MS | 6 | 7 | 24-OHCHO #<br>25-OHCHO *<br>27-OHCHO #<br>7-ketocholesterol *<br>HDL # |
| 23 | (Chen, Hsu et al. 2021) | Taiwan | MDD | HC | 40 (7/33) | 40 (15/25) | 80 (22/58) | 42.70 (13.34) | 38.80 (8.87) | Ghrelin | Serum | Immunoassay | 5 | 8,5 | Ghrelin # |
| 24 | (Xue, Zeng et al. 2020) | China | Depression | HC | 75 (27/48) | 48 (22/26) | 123 (49/74) | 26.24 (8.01) | 27.76 (8.55) | Vitamin A | Plasma | LC-MS/MS | 6 | 8 | Vitamin A * |
| 25 | (Okasha, Sabry et al. 2020) | Egypt | MDD | HC | 20 (10/10) | 20 (10/10) | 40 (20/20) | 26.6 (6.5) | 28 (2) | Vitamin D | Serum | ELISA | 4 | 10,5 | Vitamin D * |
| 26 | (Lv, Hu et al. 2020) | China | BD | HC | 28 (13/15) | 49 (20/29) | 77 (33/44) | 35.11 (15.78) | 21.12 (2.51) | MDA | Plasma | ELISA | 5 | 9 | MDA # |
| 27 | (Karadeniz, Yaman et al. 2020) | Turkey | MDD | HC | 37 (11/26) | 33 (12/21) | 70 (23/47) | 14.62 (1.60) | 14.72 (1.15) | Ghrelin, HDL | Serum | ELISA<br>Automated | 6 | 7 | Ghrelin #, HDL * |
| 28 | (Islam, Ali et al. 2020) | Bangladesh | MDD | HC | 247 (91/156) | 248 (102/146) | 495 (193/302) | 29.0 (18.4) | 34.0 (18.8) | Vitamin E | Serum | liquid-liquid extraction | 7 | 6,5 | Vitamin E # |
| 29 | (Hamed, Elmalt et al. 2020) | Egypt | MDD | HC | 60 (18/42) | 60 (22/38) | 120 (40/80) | 41.03 (4.46) | 42.8 (4.28) | MDA | Serum | Spectro. | 6 | 10 | MDA * |

| | | | | | | | | | | | | | | | |
|---|---|---|---|---|---|---|---|---|---|---|---|---|---|---|---|
| 30 | (Grudet, Wolkowitz et al. 2020) | USA | MDD | HC | 48 (21/27) | 54 (21/33) | 102 (42/60) | 39.3 (14.9) | 37.9 (13.9) | Vitamin D | Serum | LC/MS/MS | 7 | 9,5 | Vitamin D * |
| 31 | (Esnafoglu and Ozturan 2020) | Turkey | Deprrssive disorder | HC | 89 (20/69) | 43 (12/31) | 132 (32/100) | 15.08 (1.46) | 14.41 (2.32) | Vitamin D | Serum | Automated | 6 | 8 | Vitamin D * |
| 32 | (Alamolhoda , Kariman et al. 2020) | Iran | PPD | HC | 22 (0/22) | 58 (0/58) | 80 (0/80) | 27.32 (5.9) | 26 (5.09) | MDA | Serum | Atomic Absorption | 5 | 9 | MDA # |
| 33 | (Wagner, Musenbichler et al. 2019) | Germany | MDD | HC | 130 (61/69) | 61 (30/31) | 191 (91/100) | 46 (33–54) | 42 (32–55) | HDL | Serum | Automated | 7 | 7 | HDL * |
| 34 | (Su, Li et al. 2019) | China | BD | HC | 92 (42/50) | 89 (25/64) | 181 (67/114) | 24.76 (12.75) | 47.34 (13.05) | HDL | Serum | Unknown | 5 | 10 | HDL # |
| | | | UD | | 195 (49/146) | | 284 (74/210) | 44.10 (13.94) | 47.34 (13.05) | | | | | | HDL # |
| 35 | (Maes, Landucci Bonifacio et al. 2019) | Brazil | Mood Disorder | HC | 37 (30/7) | 67 (44/23) | 104 (74/30) | 42.7 (10.8) | 43.6 (11.7) | LOOH, MDA | Plasma | Spectro. | 7 | 7 | LOOH #, MDA # |
| | | | | | | | | | | | | HPLC | | | |
| 36 | (Lv, Guo et al. 2019) | China | BD | HC | 61 (34/27) | 49 (20/29) | 110 (54/56) | 27.48 (10.69) | 21.12 (2.51) | MDA | Plasma | ELISA | 5 | 9 | MDA # |
| 37 | (Hui, Yin et al. 2019) | China | BD | HC | 37 (15/22) | 37 (15/22) | 74 (30/44) | 29.78 (10.05) | 29.95 (9.02) | HDL | Serum | Automated | 5 | 8 | HDL * |
| 38 | (Hu, Wang et al. 2019) | China | BD | HC | 99 (56/43) | 99 (56/43) | 198 (112/86) | 34.00 (26.00,47.00) | 35.00 (29.00,47.00) | Apo A1, Apo B HDL | Serum | Spectro. | 6 | 10 | Apo A1 *, Apo B * HDL # |
| 39 | (Baltazar-Gaytan, Aguilar-Alonso et al. 2019) | Mexico | Depression | HC | 12 (12/0) | 12 (12/0) | 24 (24/0) | 19.08 (4.6) | 19.33 (5.1) | MDA | BrainT | Spectro. | 5 | 9 | MDA # |
| | | | | | | | | | | | | | | | MDA * |
| 40 | (Tunçel Ö, Sarısoy et al. 2018) | Turkey | BD | HC | 30 (11/19) | 30 (11/19) | 60 (22/38) | 34.4 (10.3) | 35.0 (11.3) | Ghrelin | Plasma | Automated | 7 | 9 | Ghrelin * HDL * |
| | | | | | | | | | | HDL | Serum | | | | |
| 41 | (Sowa-Kućma, Styczeń et al. 2018a) | Poland | TRD | HC | 42 (11/31) | 48 (14/34) | 90 (25/65) | 50.2 (8.5) | 45.8 (12.4) | TBARS | Serum | ELISA | 8 | 8 | TBARS # |
| | | | | | 72 (30/42) | | 120 (44/76) | 48.9 (11.9) | | | | | | | |
| 42 | (Sowa-Kućma, Styczeń et al. 2018b) | Poland | BD | HC | 133 (46/87) | 50 (14/36) | 183 (60/123) | 44.3 (12.9) | 45.8 (12.4) | TBARS | Serum | ELISA | 8 | 8 | TBARS # |
| | | | MDD | | 114 (41/73) | | 164 (55/109) | 49.4 (10.7) | | | | | | | |
| 43 | (Segoviano-Mendoza, Cárdenas-de la Cruz et al. 2018) | Mexico | MDD | HC | 204 (35/169) | 206 (40/166) | 410 (75/335) | 37.3 (10.0) | 36.8 (6.6) | HDL | Serum | Automated | 7 | 8 | HDL * |
| | | | | | 59 (17/42) | | 265 (57/208) | 35.2 (10.5) | | | | | | | |
| 44 | (Ramachandran Pillai, Wilson et al. 2018) | India | PPD | HC | 186 (0/186) | 250 (0/250) | 436 (0/436) | 25 (23-29) | 24 (22-28) | HDL | Serum | Automated | 7 | 7,5 | HDL # |

| | | | | | | | | | | | | | | | |
|---|---|---|---|---|---|---|---|---|---|---|---|---|---|---|---|
| 45 | (Petrov, Aldoori et al. 2018) | USA | BD<br>MDD | HC | 13 (13/0)<br>11 (11/0) | 12 (12/0) | 25 (25/0)<br>23 (23/0) | 13.9 (2.02)<br>14.09 (1.22) | 14 (2.42) | Vitamin D | Serum | HPLC | 5 | 9 | Vitamin D #<br>Vitamin D * |
| 46 | (Jordan, Dobrowolny et al. 2018) | Germany | MDD | HC | 18 (11/7) | 43 (26/17) | 61 (37/24) | 43.9 (15.4) | 35.3 (14.5) | MDA | Plasma | HPLC | 7 | 7 | MDA # |
| 47 | (Islam, Islam et al. 2018) | Bangladesh | MDD | HC | 247 (91/156) | 248 (102/146) | 495 (193/302) | 33.03 (10.89) | 33.55 (9.58) | MDA | Serum | ELISA | 7 | 5 | MDA # |
| 48 | (Enko, Brandmayr et al. 2018) | Austria | Depression | HC | - | - | 0 (0/0) | - | - | HDL | Plasma | Automated | 5 | 12 | HDL # |
| 49 | (Diniz, Mendes-Silva et al. 2018) | Brazil | Depression | HC | 76 (10/66) | 47 (3/44) | 123 (13/110) | 72.6 (7.7) | 70.1 (7.1) | 8-Isoprostane<br>TBARS | Plasma | ELISA<br>Spectro. | 6 | 9 | 8-Isoprostane #<br>TBARS # |
| 50 | (Ceylan, Tuna et al. 2018) | Turkey | BD | HC | 32 (12/20) | 51 (21/30) | 83 (33/50) | 37.63 (9.96) | 36.28 (11.45) | MDA | Plasma | HPLC | 6 | 10,5 | MDA * |
| 51 | (Baghai, Varallo-Bedarida et al. 2018) | Germany | MDD | HC | 100 (37/63) | 104 (47/57) | 204 (84/120) | 46.6 (14.8) | 54.7 (14.4) | HDL | Unknown | Unknown | 5 | 9,5 | HDL * |
| 52 | (Altunsoy, Yüksel et al. 2018) | Turkey | BD | HC | 26 (14/12)<br>31 (16/15) | 40 (23/17) | 66 (37/29)<br>71 (39/32) | 33.27 (12.09)<br>35.90 (8.84) | 37.58 (9.59) | Vitamin D | Serum | Automated | 6 | 8 | Vitamin D * |
| 53 | (Algul and Ozcelik 2018) | Turkey | MDD | HC | 30 (13/17) | 30 (15/15) | 60 (28/32) | 38.0 (16)<br>37.1 (14) | 36.2 (10) | Ghrelin | Plasma | ELISA | 8 | 6,5 | Ghrelin # |
| 54 | (Abedi, Bovayri et al. 2018) | Iran | PPD | HC | 60 (0/60) | 60 (0/60) | 120 (0/120) | 26.43 (4.27) | 27.6 (4.73) | Vitamin D | Serum | ELISA | 6 | 7,5 | Vitamin D * |
| 55 | (Newton, Naiberg et al. 2017) | Canada | BD | HC | 29 (12/17) | 25 (12/13) | 54 (24/30) | 16.80 (1.86) | 15.4 (1.68) | 4-HNE<br>LOOH | Serum | ELISA<br>Spectro. | 4 | 10 | 4-HNE *<br>LOOH * |
| 56 | (Moreira, Jansen et al. 2017) | Brazil | MDD | HC | 77 (13/64)<br>32 (6/26) | 863 (383/480) | 940 (396/544)<br>895 (389/506) | 26.09 (2.22)<br>25.59 (2.24) | 25.81 (2.17) | HDL | Serum | Unknown | 6 | 8 | HDL * |
| 57 | (Messaoud, Mensi et al. 2017) | Tunisia | MDD | HC | 110 (35/75)<br>52 (19/33) | 151 (50/101) | 261 (85/176)<br>203 (69/134) | 44.33 (10.50)<br>29.84 (8.78) | 38.92 (13.28) | HDL | Plasma | Automated | 7 | 7,5 | HDL * |
| 58 | (Lindqvist, Dhabhar et al. 2017) | USA | MDD | HC | 50 (23/27) | 55 (22/33) | 105 (45/60) | 39.6 (14.7) | 37.6 (13.9) | 8-Isoprostane | Plasma | GC–MS | 5 | 9,5 | 8-Isoprostane # |
| 59 | (Chowdhury, Hasan et al. 2017) | Bangladesh | BD | HC | 60 (40/20) | 60 (34/26) | 120 (74/46) | 26.27 (1.06) | 25.73 (0.81) | MDA<br>Vitamin A<br>Vitamin E | Serum | Spectro.<br>RP-HPLC | 5 | 8,5 | MDA #<br>Vitamin A *<br>Vitamin E * |

| | | | | | | | | | | | | | | | |
|---|---|---|---|---|---|---|---|---|---|---|---|---|---|---|---|
| 60 | (Camkurt, Fındıklı et al. 2017) | Turkey | MDD | HC | 33 (0/33) | 37 (0/37) | 70 (0/70) | 34.6 (12.4) | 33.84 (10.7) | MDA | Serum | Spectro. | 4 | 11 | MDA # |
| 61 | (Akgün, Köken et al. 2017) | Turkey | BD | HC | 20 (6/14)<br>20 (5/15) | 22 (6/16) | 42 (12/30)<br>42 (11/31) | 39.8 (7.5)<br>46.4 (10.2) | 31.3 (3.7) | MDA | Serum | HPLC | 7 | 10 | MDA # |
| 62 | (Tunçel Ö, Akbaş et al. 2016) | Turkey | BD | HC | 30 (7/23) | 30 (7/23) | 60 (14/46) | 14.7 (2.03) | 14.7 (2.03) | Ghrelin | Serum | ELISA | 7 | 8,5 | Ghrelin # |
| 63 | (Tsai and Huang 2016) | Taiwan | MDD | HC | 21 (4/17) | 40 (10/30) | 61 (14/47) | 49.6 (7.0) | 33.0 (5.7) | TBARS | Serum | ELISA | 6 | 6,5 | TBARS # |
| 64 | (Siwek, Sowa-Kucma et al. 2016) | Poland | BD | HC | 129 (49/80) | 50 (14/36) | 179 (63/116) | 44 (13) | 46 (12) | TBARS | Serum | Spectro. | 7 | 8 | TBARS # |
| 65 | (Scola, McNamara et al. 2016) | USA | BD | HC | 16 (6/10) | 13 (7/6) | 29 (13/16) | 15.5 (2.4) | 17.8 (2.5) | 4-HNE<br>8-Isoprostane<br>LOOH | Serum | ELISA<br>ELISA<br>Spectro. | 7 | 5,5 | 4-HNE #<br>8-Isoprostane #<br>LOOH * |
| 66 | (Elvsåshagen, Zuzarte et al. 2016) | Norway | BD | HC | 29 (9/20) | 33 (15/18) | 62 (24/38) | 35.0 (6.9) | 34.4 (9.5) | 4-HNE<br>LOOH | Blood Cells | ELISA<br>Spectro. | 5 | 9 | 4-HNE #<br>LOOH # |
| 67 | (Camkurt, Fındıklı et al. 2016) | Turkey | MDD | HC | 50 (16/34) | 50 (22/28) | 100 (38/62) | 34.6 (12.4) | 33.84 (10.7) | MDA | Plasma | Spectro. | 5 | 7,5 | MDA # |
| 68 | (Mondin, de Azevedo Cardoso et al. 2016) | Brazil | BD<br>MDD | HC | 48 (12/36)<br>73 (18/55) | 94 (40/54) | 142 (52/90)<br>167 (58/109) | 21.92 (2.32) | 22.40 (2.25) | TBARS | Serum | Spectro. | 4 | 15 | TBARS ND |
| 69 | (Tunçel, Sarısoy et al. 2015) | Turkey | MDD | HC | 18 (8/10) | 18 (8/10) | 36 (16/20) | 32.6 (9.8) | 34.8 (11.3) | MDA | Serum | Spectro. | 5 | 7,5 | MDA # |
| 70 | (Tsai and Huang 2015) | Taiwan | MDD | HC | 23 (13/10) | 40 (20/20) | 63 (33/30) | 41.3 (12.0) | 30.4 (5.0) | TBARS | Unknown | Spectro. | 6 | 6,5 | TBARS # |
| 71 | (Sarandol, Sarandol et al. 2015) | Turkey | BD | HC | 26 (10/16) | 25 (10/15) | 51 (20/31) | 25.6 (7.0) | 23.5 (9.2) | Apo A1<br>Apo B<br>HDL<br>MDA<br>MDA<br>PON1<br>Vitamin E | Serum<br>Serum<br>Serum<br>Plasma<br>Blood Cells<br>Serum<br>Plasma | Automated<br>Automated<br>Automated<br>HPLC<br>chemically<br>Spectro.<br>HPLC | 6 | 9 | Apo A1 *<br>Apo B *<br>HDL *<br>MDA #<br>MDA #<br>PON1 #<br>Vitamin E * |
| 72 | (Rosso, Cattaneo et al. 2015) | Italy | BD | HC | 57 (18/39) | 49 (25/24) | 106 (43/63) | 50.2 (14.2) | 32 (5.14) | Ghrelin | Serum | multiplex | 6 | 7 | Ghrelin * |
| 73 | (Ormonde do Carmo, Mendes-Ribeiro et al. 2015) | Brazil | MDD | HC | 22 (4/18) | 27 (10/17) | 49 (14/35) | 31 (2) | 33 (2) | HDL | Unknown | Automated | 6 | 5 | HDL # |

| | | | | | | | | | | | | | | | |
|---|---|---|---|---|---|---|---|---|---|---|---|---|---|---|---|
| 74 | (Kaufmann, Gazal et al. 2015) | Brazil | MDD | HC | 49 (10/39) | 49 (10/39) | 98 (20/78) | 24.06 (3.52) | 24.14 (3.22) | TBARS | Serum | Spectro. | 5 | 9 | TBARS # |
| 75 | (Kahl, Schweiger et al. 2015) | Germany | MDD | HC | 10 (3/7) | 90 (57/33) | 100 (60/40) | 51.3 (14.7) | 52.3 (14.9) | HDL | Plasma | Automated | 5 | 9,5 | HDL * |
| 76 | (Cimen, Gumus et al. 2015) | Turkey | MDD | HC | 18 (7/11) | 18 (8/10) | 36 (15/21) | 42.17 (10.16) | 37.89 (10.62) | MDA | Plasma | Spectro. | 6 | 5,5 | MDA #, MDA * |
| 77 | (Bengesser, Lackner et al. 2015) | Austria | BD | HC | 113 (63/50) | 78 (30/48) | 191 (93/98) | 45.2 (14.3) | 42.1 (17.0) | MDA<br>TBARS | Serum | HPLC | 5 | 10,5 | MDA*<br>TBARS # |
| 78 | (Andreazza, Gildengers et al. 2015) | Canada | BD | HC | 110 (24/86) | 75 (35/40) | 185 (59/126) | 63.86 (9.70) | 66.00 (9.62) | 4-HNE<br>LOOH | Plasma | Spectro. | 7 | 8 | 4-HNE #<br>LOOH # |
| 79 | (Versace, Andreazza et al. 2014) | Canada | BD | HC | 24 (8/16) | 19 (9/10) | 43 (17/26) | 33.2 (7.7) | 34.0 (4.4) | 4-HNE<br>LOOH | Serum | ELISA<br>Spectro. | 5 | 9 | 4-HNE *<br>LOOH # |
| 80 | (Vargas, Nunes et al. 2014) | Brazil | BD<br>MDD | HC | 49 (12/37)<br>92 (22/70) | 201 (82/119) | 250 (94/156)<br>293 (104/189) | - | - | HDL | Serum | Automated | 6 | 7 | HDL * |
| 81 | (Spanemberg, Caldieraro et al. 2014) | Brazil | MDD | HC | 20 (2/18)<br>13 (3/10) | 54 (14/40) | 74 (16/58)<br>67 (17/50) | 48.4 (7.7)<br>52.8 (10.7) | 47.4 (9.97) | TBARS | Serum | Spectro. | 5 | 8,5 | TBARS #,<br>TBARS * |
| 82 | (Patra, Khandelwal et al. 2014) | India | MDD | HC | 30 (19/11) | 30 (19/11) | 60 (38/22) | 32.47 (11.41) | 32.63 (11.20) | HDL | Serum | Spectro. | 6 | 7,5 | HDL # |
| 83 | (Ozsoy, Besirli et al. 2014) | Turkey | Depression | HC | 28 (11/17) | 21 (10/11) | 49 (21/28) | 48.00 (14.66) | 40.86 (12.39) | Ghrelin | Serum | Immunoassay | 5 | 9 | Ghrelin # |
| 84 | (Milaneschi, Hoogendijk et al. 2014) | Netherlands | Depressive disorders | HC | 790 (242/548)<br>1102 (362/740) | 494 (194/300) | 1284 (436/848)<br>1596 (556/1040) | 43.4 (12.5)<br>40.9 (12.1) | 40.1 (14.9) | Vitamin D | Serum | ID-XLC-MS | 7 | 7,5 | Vitamin D # |
| 85 | (Józefowicz, Rabe-Jabłońska et al. 2014) | Poland | MDD | HC | 20 (8/12) | 40 (18/22) | 60 (26/34) | 38.6 (3.3) | 39.8 (2.0) | Ghrelin<br>HDL | Serum | ELISA<br>Automated | 8 | 6,5 | Ghrelin #<br>HDL * |
| 86 | (de Sousa, Zarate et al. 2014) | Brazil | BD | HC | 29 (8/21) | 28 (12/16) | 57 (20/37) | 28.4 (5.5) | 28.0 (7.2) | TBARS | Plasma | Spectro. | 6 | 5,5 | TBARS * |
| 87 | (Cudney, Sassi et al. 2014) | Canada | BD | HC | 52 (0/52) | 30 (0/30) | 82 (0/82) | 40.75 (12.48) | 35.93 (11.71) | MDA | Serum | Spectro. | 5 | 9 | MDA * |
| 88 | (Bortolasci, Vargas et al. 2014) | Brazil | BD<br>MDD | HC | 45 (12/33)<br>91 (22/69) | 199 (82/117) | 244 (94/150)<br>290 (104/186) | 44.5 (8.9)<br>47.0 (8.2) | 46.4 (8.3) | HDL<br>PON1 | Serum | Spectro. | 7 | 7,5 | HDL #<br>PON1 #<br>HDL #<br>PON1 # |

| | | | | | | | | | | | | | | | |
|---|---|---|---|---|---|---|---|---|---|---|---|---|---|---|---|
| 89 | (Bajpai, Verma et al. 2014) | India | UP | HC | 60 (25/35) | 40 (40/0) | 100 (65/35) | - | - | MDA | Serum | Spectro. | 6 | 8 | MDA # |
| 90 | (Aydemir, Çubukçuoğlu et al. 2014) | Turkey | BD | HC | 51 (27/24) | 50 (27/23) | 101 (54/47) | 40.8 (11.5) | 39.8 (11.2) | MDA | Plasma | Spectro. | 6 | 8 | MDA # |
| 91 | (Vargas, Nunes et al. 2013) | Brazil | BD | HC | 49 (12/37) | 201 (82/119) | 250 (94/156) | - | - | HDL | Serum | Automated | 6 | 7 | HDL * |
| | | | MDD | | 92 (22/70) | 201 (82/119) | 293 (104/189) | - | - | | | | | | |
| 92 | (Rybka, Kędziora-Kornatowska et al. 2013) | Poland | Depressive disorders | HC | 15 (15/0) | 19 (19/0) | 34 (34/0) | 59.7 (1.91) | 62.3 (2.84) | MDA | Blood Cells | Spectro. | 6 | 6,5 | MDA # |
| 93 | (Rawdin, Mellon et al. 2013) | USA | MDD | HC | 19 (7/12) | 20 (7/13) | 39 (14/25) | 37.00 (10.77) | 36.42 (12.13) | 8-Isoprostane | Plasma | GC–MS | 7 | 7,5 | 8-Isoprostane* |
| 94 | (Maes, Kubera et al. 2013) | Belgium | MDD | HC | 113 (54/59) | 28 (9/19) | 141 (63/78) | 43.5 (11.3) | 40.3 (11.6) | OxLDL<br>Peroxide | Serum | ELISA | 8 | 7,5 | OxLDL #<br>Peroxide # |
| 95 | (Jamilian, Bagherzadeh et al. 2013) | Iran | Depression | HC | 100 (35/65) | 100 (50/50) | 200 (85/115) | 35.84 (8.20) | 35.26 (4.82) | Vitamin D | Serum | Unknown | 7 | 7,5 | Vitamin D * |
| 96 | (Gubert, Stertz et al. 2013) | Brazil | BD | HC | 12 (8/4) | 12 (8/4) | 24 (16/8) | 41.83 (4.1) | 40.55 (4.3) | MDA | Plasma | Spectro. | 6 | 7,5 | MDA # |
| 97 | (Khanzode, Dakhale et al. 2003) | India | MDD | HC | 62 (28/34) | 40 (22/18) | 102 (50/52) | 43.77 (12.85) | 40.85 (10.22) | MDA | Serum | Spectro. | 6 | 10 | MDA # |
| 98 | (Erman, Erman et al. 2013) | Turkey | MDD | HC | 20 (8/12) | 40 (18/22) | 60 (26/34) | 38.6 (3.3) | 39.8 (2.0) | Ghrelin<br>HDL | Serum | ELISA<br>Automated | 8 | 6,5 | Ghrelin #<br>HDL * |
| 99 | (Baek and Park 2013) | South Korea | Depression | HC | 80 (21/59) | 80 (28/52) | 160 (49/111) | 44.85 (1.77) | 44.47 (1.63) | TBARS | Serum | Spectro. | 6 | 7 | TBARS # |
| 100 | (Vuksan-Cusa, Sagud et al. 2013) | Croatia | BD | HC | 60 (31/29) | 59 (30/29) | 119 (61/58) | 44.4 (15.8) | 42.2 (8.7) | HDL | Serum | Automated | | | HDL ND |
| 101 | (Andreazza, Wang et al. 2013) | Canada | BD | HC | 16 (3/13) | 26 (15/11) | 42 (18/24) | 58 (5.51) | 60 (2.79) | 4-HNE<br>8-Isoprostane<br>LOOH | BrainT | EIAs<br>ELISA<br>Spectro. | 6 | 8 | 4-HNE #<br>8-Isoprostane #<br>LOOH * |
| 102 | (Stefanescu and Ciobica 2012) | Romania | Depression | HC | 15 (6/9) | 20 (7/13) | 35 (13/22) | 44.66 (6.8) | 46.3 (7.8) | MDA | Serum | Spectro. | 6 | 7,5 | MDA # |
| | | | | | 16 (5/11) | | | 50.44 (8.9) | | | | | | | |
| 103 | (Pomara, Bruno et al. 2012) | USA | Depression | HC | 28 (18/10) | 19 (12/7) | 47 (30/17) | 66.5 (5.4) | 68.1 (7.3) | 8-Isoprostane | CSF | ELISA | 5 | 8 | 8-Isoprostane # |
| 104 | (Magalhães, Jansen et al. 2012) | Brazil | BD | HC | 55 (15/40) | 94 (40/54) | 149 (55/94) | 21.7 (0.31) | 22.4 (0.23) | TBARS | Serum | Spectro. | 7 | 7,5 | TBARS #,<br>TBARS * |
| | | | MDD | | 82 (19/63) | | | 21.8 (0.22) | | | | | | | |

| | | | | | | | | | | | | | | | |
|---|---|---|---|---|---|---|---|---|---|---|---|---|---|---|---|
| 105 | (Matsuo, Nakano et al. 2012) | Japan | MDD | HC | 24 (13/11) | 24 (10/14) | 48 (23/25) | 50.7 (12.5) | 54.4 (17.2) | Ghrelin | Plasma | ELISA | 4 | 9 | Ghrelin # |
| 106 | (Ishitobi, Kohno et al. 2012) | Japan | MDD | HC | 38 (16/22)<br>30 (15/15) | 103 (59/44) | 141 (75/66) | 38.2 (13.1)<br>39.6 (11.6) | 32.7 (5.1) | Ghrelin | Serum | ELISA | 7 | 9 | Ghrelin # |
| 107 | (Hocaoglu, Kural et al. 2012) | Turkey | MDD | HC | 30 (6/24) | 30 (16/14) | 60 (22/38) | 38 (13) | 30 (9) | HDL | Serum | Automated | 4 | 10 | HDL # |
| 108 | (Ghodake, Suryakar et al. 2012) | India | MDD | HC | 30 (0/0) | 30 (0/0) | 60 (0/0) | 32.20 (6.4) | - | MDA<br>Vitamin E | Plasma | Spectro. | 6 | 8 | MDA #<br>Vitamin E * |
| 109 | (Ezzaher, Haj Mouhamed et al. 2012) | Tunisia | BD | HC | 90 (59/31) | 92 (60/32) | 182 (119/63) | 37.2 (11.8) | 34.1 (14.0) | TBARS | Plasma | Spectro. | 5 | 10 | TBARS # |
| 110 | (Banerjee, Dasgupta et al. 2012) | India | BD | HC | 48 (33/15)<br>25 (18/7) | 35 (26/9) | 83 (59/24) | 39.40 (12.6)<br>33.68 (7.56) | 35.88 (9.05) | TBARS | Serum | Spectro. | 6 | 6,5 | TBARS # |
| 11 | (Bal, Acar et al. 2012) | Turkey | MDD | HC | 42 (5/37) | 38 (6/32) | 80 (11/69) | 44.1 (12.3) | 44.0 (9.3) | MDA<br>Vitamin E | Plasma | Spectro. | 6 | 8 | MDA #<br>Vitamin E * |
| 112 | (Vila-Rodriguez, Honer et al. 2011) | UK | BD | HC | 34 (16/18) | 35 (26/9) | 69 (42/27) | 45.4 (10.7) | 44.2 (7.6) | Apo E | BrainT | ELISA | 5 | 10 | Apo E *, Apo E # |
| 113 | (Sonal Sukreet and Chaturvedi 2011) | India | BD | HC | 100 (58/42) | 52 (26/26) | 152 (84/68) | 63 (12) | 52 (12) | HDL<br>MDA | Serum | Automated<br>Spectro. | 6 | 10 | HDL #<br>MDA # |
| 114 | (Sadeghi, Roohafza et al. 2011) | Iran | MDD | HC | 153 (90/63) | 147 (69/78) | 300 (159/141) | 31.21 (10.41) | 32.00 (8.21) | Apo A1<br>Apo B<br>HDL | Serum | Immunoassay<br>Immunoassay<br>direct method | 5 | 10,5 | Apo A1 *<br>Apo B #<br>HDL # |
| 115 | (Murat Can 2011) | Turkey | BD<br>MDD | HC | 39 (20/19)<br>32 (11/21) | 30 (15/15) | 69 (35/34) | 38.3 (13.4)<br>35.5 (13.7) | 36.1 (12.2) | MDA | Serum | Spectro. | 5 | 11 | MDA # |
| 116 | (Kotan, Sarandol et al. 2011) | Turkey | MDD | HC | 50 (11/39) | 44 (10/34) | 94 (21/73) | 33.1 (10.0) | 33.2 (7.9) | Apo A1<br>Apo B<br>HDL<br>MDA<br>OxLDL<br>PON1<br>Vitamin A<br>Vitamin E | Serum<br>Serum<br>Serum<br>Plasma<br>Serum<br>Serum<br>Plasma<br>Plasma | Immunoassay<br>Immunoassay<br>Spectro.<br>HPLC<br>ELISA<br>Spectro.<br>Spectro.<br>Spectro. | 7 | 6 | Apo A1 #<br>Apo B *<br>HDL *<br>MDA #<br>OxLDL *<br>PON1 *<br>Vitamin A *<br>Vitamin E # |
| 117 | (Kapczinski, Dal-Pizzol et al. 2011) | Brazil | BD<br>Depression | HC | 20 (8/12)<br>20 (4/16) | 80 (32/48) | 100 (40/60) | 37.9 (12.1)<br>46.1 (9.3) | 40.7 (12.5) | TBARS | Serum | Spectro. | 5 | 10 | TBARS #<br>TBARS * |
| 118 | (Yager, Forlenza et al. 2010) | USA | Depression | HC | 73 (12/61) | 72 (13/59) | 145 (25/120) | 28.4 (9.1) | 28.8 (9.2) | 8-Isoprostane | Serum | ELISA | 7 | 5,5 | 8-Isoprostane # |
| 119 | | Netherlands | MDD | HC | 761 (256/505) | 629 (243/386) | | 41.7 (12.9) | 41.2 (12.9) | HDL | Serum | Automated | 6 | 8,5 | HDL * |

| | | | | | | | | | | | | | | | |
|---|---|---|---|---|---|---|---|---|---|---|---|---|---|---|---|
| | (van Reedt Dortland, Giltay et al. 2010) | | | | 1071 (316/755) | | 1390 (499/891) | 42.4 (12.9) | | | | | | | |
| 120 | (S. Nur Aksoy and Abdurrahman Altındag 2010) | Turkey | BD | HC | 30 (12/18) | 30 (13/17) | 60 (25/35) | 32.8 (1.58) | 34.2 (1.88) | MDA | Plasma | Spectro. | 5 | 8,5 | MDA # |
| 121 | (Maes, Mihaylova et al. 2010) | Belgium | MDD | HC | 54 (23/31) | 37 (12/25) | 91 (35/56) | 43.5 (11.6) | 43.6 (11.1) | OxLDL<br>Peroxide | Serum | ELISA | 7 | 7,5 | OxLDL #<br>Peroxide # |
| 122 | (Lehto, Niskanen et al. 2010) | Finland | MDD | HC | 43 (19/24)<br>45 (20/25) | 88 (39/49) | 131 (58/73) | 50.56 (4.80)<br>49.09 (6.63) | 49.86 (8.01) | HDL | Serum | Automated | 6 | 9 | HDL * |
| 123 | (Wang, Shao et al. 2009) | Canada | BD<br>MDD | HC | 15 (/) | 15 (/) | 30 (0/0) | - | - | 4-HNE | BrainT | Immunoassay | 4 | 12 | 4-HNE # |
| 124 | (Sagud, Mihaljevic-Peles et al. 2009) | Croatia | BD<br>MDD | HC | 22 (0/22)<br>19 (0/19)<br>34 (0/34) | 50 (0/50) | 72 (0/72) | 48.5 (13.2)<br>44.1 (12.9)<br>50.1 (6.6) | 44.7 (12.8) | HDL | Serum | Automated | 5 | 7,5 | HDL * |
| 125 | (Maes, Mihaylova et al. 2009) | Belgium | Depression | HC | 22 (5/17) | 35 (15/20) | 57 (20/37) | 42.1 (10.5) | 45.4 (10.1) | CoEQ10 | Plasma | HPLC | 5 | 8,5 | CoEQ10 * |
| 125 | (Kodydková, Vávrová et al. 2009) | Czech Republic | Depression | HC | 35 (0/35) | 35 (0/35) | 70 (0/70) | 64.5 (50.0–75.1) | 65.0 (53.2–77.0) | Apo A1<br>Apo B<br>HDL<br>PON1 | Serum | Automated<br>Automated<br>Automated<br>Spectro. | 6 | 8,5 | Apo A1 *<br>Apo B #<br>HDL *<br>PON1 * |
| 127 | (Kluge, Schussler et al. 2009) | Germany | MDD | HC | 11 (0/11)<br>9 (9/0) | 11 (0/11)<br>9 (9/0) | 22 (0/22)<br>18 (18/0) | 39.4 (10.2)<br>38.3 (10.4) | 38.7 (10.8)<br>39.1 (11.2) | Ghrelin | Plasma | Immunoassay | 7 | 5,5 | Ghrelin # |
| 128 | (Gałecki, Szemraj et al. 2009) | Poland | MDD | HC | 50 (22/28) | 30 (14/16) | 80 (36/44) | 36.7 (5.2) | 32.1 (4.3) | MDA | Blood Cells | Spectro. | 6 | 6 | MDA # |
| 129 | (Barim, Aydin et al. 2009) | Turkey | Depression | HC | 24 (12/12) | 22 (11/11) | 46 (23/23) | 35.7 (11.2) | 36.1 (9.3) | Ghrelin<br>HDL<br>PON1 | Plasma | Spectro.<br>Automated<br>Spectro. | 6 | 5,5 | Ghrelin *<br>HDL *<br>PON1 * |
| 130 | (Lehto, Hintikka et al. 2008) | Finland | MDD | HC | 63 (19/44) | 61 (17/44) | 124 (36/88) | 54.68 (8.59) | 55.59 (8.96) | HDL | Serum | Automated | 6 | 9,5 | HDL * |
| 131 | (Kurt, Güler et al. 2008) | Turkey | MDD | HC | 36 (11/25) | 25 (8/17) | 61 (19/42) | 31.14 (11.98) | 32.24 (9.40) | Ghrelin | Serum | ELISA | 6 | 7,5 | Ghrelin # |
| 132 | (Kunz, Gama et al. 2008) | Brazil | BD | HC | 32 (12/20)<br>19 (3/16)<br>32 (18/14) | 32 (11/21) | 64 (23/41) | 40.28 (12.00)<br>43.43 (8.00)<br>40.13 (12.65) | 40.69 (12.12) | TBARS | Serum | Spectro. | 5 | 10 | TBARS # |
| 133 | (Dimopoulos, Piperi et al. 2008) | Greece | Depression | HC | 33 (13/20) | 33 (13/20) | 66 (26/40) | 65.81 (3.36) | 65.39 (4.06) | 8-Isoprostane | Serum | Immunoassay | 5 | 10,5 | 8-Isoprostane # |
| 134 | (Dean, Digney et al. 2008) | Australia | BD | HC | 8 (6/2) | 23 (14/9) | 31 (20/11) | 38.3 (10.4) | 29.0 (7.0) | Apo E | Plasma | Western blot | 5 | 8 | Apo E * |

| | | | | | | | | | | | | | | | |
|---|---|---|---|---|---|---|---|---|---|---|---|---|---|---|---|
| 135 | (Sarandol, Sarandol et al. 2007) | Turkey | MDD | HC | 96 (24/72) | 54 (14/40) | 150 (38/112) | 40 (11) | 37 (9) | MDA<br>MDA<br>Vitamin E | Plasma<br>Blood Cells<br>Serum | HPLC<br>Spectro.<br>HPLC | 7 | 7,5 | MDA #<br>Vitamin E # |
| 136 | (Machado-Vieira, Andreazza et al. 2007) | Brazil | BD | HC | 21 (16/5)<br>32 (14/18)<br>32 (20/12) | 32 (21/11) | 53 (37/16) | 43.4 (8.0)<br>40.1 (12.6)<br>40.3 (11.3) | 40.7 (12.1) | TBARS | Plasma | Spectro. | 6 | 6,5 | TBARS # |
| 137 | (Kurt, Guler et al. 2007) | Turkey | BD<br>MDE | HC | - | - | 0 (0/0) | 0 (0) | - | Ghrelin | Serum | ELISA | 6 | 7,5 | Ghrelin # |
| 138 | (Andreazza, Cassini et al. 2007) | Brazil | BD | HC | 21 (16/5)<br>32 (14/18)<br>32 (20/12) | 32 (21/11) | 53 (37/16)<br>64 (35/29)<br>64 (41/23) | 43.4 (8.0)<br>40.1 (12.6)<br>40.3 (11.3) | 40.7 (12.1) | TBARS | Serum | Spectro. | 8 | 8 | TBARS # |
| 139 | (Sarandol, Sarandol et al. 2006) | Turkey | MDD | HC | 86 (24/62) | 36 (10/26) | 122 (34/88) | 40.5 (10.5) | 37.2 (7.3) | Apo A1<br>Apo B<br>HDL<br>MDA<br>PON1 | Plasma<br>Plasma<br>Serum<br>Serum<br>Serum | Automated | 7 | 8 | Apo A1 *<br>Apo B #<br>HDL #<br>MDA #<br>PON1 * |
| 140 | (Huang 2005) | Taiwan | MDD | HC | 109 (32/77) | 59 (22/37) | 168 (54/114) | 31.4 (8.5) | 29.5 (4.4) | HDL | Serum | Automated | 5 | 10 | HDL * |
| 141 | (Hamidreza Roohafza 2005) | Iran | MDD | HC | 25 (5/20) | 25 (7/18) | 50 (12/38) | 33.2 (6.1) | 33.2 (7.7) | HDL | Serum | Spectro. | 6 | 7 | HDL * |
| 142 | (Digney, Keriakous et al. 2005) | Australia | BD | HC | 8 (4/4) | 17 (9/8) | 25 (13/12) | 59 (3.6) | 47 (14.018) | Apo E | BrainT | Spectro. | 5 | 10 | Apo E # |
| 143 | (Selley 2004) | Australia | Depression | HC | 25 (13/12) | 25 (12/13) | 50 (25/25) | 46.1 (12.4) | 47.0 (10.0) | 4-HNE | Plasma | GC–MS | 5 | 10 | 4-HNE # |
| 144 | (Ozcan, Gulec et al. 2004) | Turkey | Affective disorder | HC | 30 (16/14) | 21 (11/10) | 51 (27/24) | 39.8 (13.1) | 39.8 (13.1) | MDA | Blood Cells | Spectro. | 4 | 10 | MDA # |
| 145 | (Karlović, Buljan et al. 2004) | Croatia | MDD | HC | 38 (38/0) | 39 (39/0) | 77 (77/0) | 46.2 (11.3) | 43.8 (10.1) | HDL | Serum | Automated | 6 | 7 | HDL * |
| 146 | (Huang and Chen 2004) | Taiwan | MDD | HC | 68 (31/37) | 39 (20/19) | 107 (51/56) | 43.5 (14.0) | 50.2 (11.2) | HDL | Serum | Automated | 5 | 10 | HDL # |
| 147 | (Eskandari, Martinez et al. 2007) | USA | PPD | HC | 89 (0/89) | 44 (0/44) | 133 (0/133) | 35 (6.9) | 35 (6.8) | Vitamin D | Serum | Immunoassay | 5 | 10 | Vitamin D * |
| 148 | (Ranjekar, Hinge et al. 2003) | India | BD | HC | 10 (10/0) | 31 (31/0) | 41 (41/0) | 40.8 (8.29) | 40.9 (8.84) | Apo A1<br>Apo B<br>HDL<br>TBARS | Plasma<br>Plasma<br>Plasma<br>Plasma | Automated<br>Automated<br>Automated<br>Spectro. | 6 | 8 | Apo A1 #<br>Apo B #<br>HDL *<br>TBARS # |
| 149 | (Huang, Wu et al. 2003) | Taiwan | MDD | HC | 68 (31/37) | 39 (20/19) | 107 (51/56) | 43.5 (14.0) | 50.2 (11.2) | HDL | Serum | Automated | 5 | 10 | HDL # |

| | | | | | | | | | | | | | | | |
|---|---|---|---|---|---|---|---|---|---|---|---|---|---|---|---|
| 150 | (Kuloglu, Ustundag et al. 2002) | Turkey | BD | HC | 23 (/) | 20 (/) | 43 (0/0) | - | - | MDA | Plasma | Spectro. | 4 | 12 | MDA # |
| 151 | (Sevincok, Buyukozturk et al. 2001) | Turkey | MDD | HC | 27 (7/20) | 24 (6/18) | 51 (13/38) | 33.29 (6.12) | 33.20 (7.78) | HDL | Serum | Automated | 6 | 7 | HDL * |
| 152 | (Bilici, Efe et al. 2001) | Turkey | MDD | HC | 18 (5/13)<br>12 (4/8) | 32 (16/16) | 50 (21/29) | 42.2 (9.7)<br>40.4 (6.4) | 42.1 (7.4) | MDA | Plasma | Spectro. | 6 | 8,5 | MDA # |
| 153 | (Schneider, Weber et al. 2000) | Germany | MDD | HC | 25 (11/14) | 31 (19/12) | 56 (30/26) | 57.6 (3.5) | 38.8 (3.2) | Vitamin D | Plasma | Immunoassay | 5 | 10 | Vitamin D * |
| 154 | (Maes, De Vos et al. 2000) | Belgium | MDD | HC | 42 (12/30) | 26 (11/15) | 68 (23/45) | 54.0 (13.6) | 49.7 (8.7) | Vitamin E | Serum | Unknown | 4 | 8,5 | Vitamin E * |
| 155 | (Fadillioğlu, Kaya et al. 2000) | Turkey | Depression | HC | 8 (8/0) | 8 (8/0) | 16 (16/0) | 20.3 (0.6) | 20.8 (0.4) | MDA | Plasma, Blood Cells | Spectro. | 4 | 10,5 | MDA *, MDA# |
| 156 | (Maes, Christophe et al. 1999) | Belgium | MDD | HC | 34 (18/16) | 14 (9/5) | 48 (27/21) | 52.2 (13.6) | 48.3 (15.2) | LCAT | Serum | Spectro. | 6 | 8 | LCAT * |
| 157 | (Maes, Smith et al. 1997) | Belgium | MDD | HC | 36 (5/31) | 28 (5/23) | 64 (10/54) | 51.1 (13.7) | 47.7 (14.2) | HDL | Serum | Automated | 5 | 8,5 | HDL * |
| 158 | (Olusi and Fido 1996) | Kuwait | MDD | HC | 100 (64/36) | 100 (64/36) | 200 (128/72) | 39.58 (10.2) | 39.96 (9.8) | Apo A1<br>Apo B<br>HDL | Serum | Automated | 6 | 8 | Apo A1 #<br>Apo B *<br>HDL * |
| 159 | (Maes, Delanghe et al. 1994) | Belgium | MDD | HC | 16 (16/0)<br>14 (14/0)<br>17 (17/0) | 26 (26/0) | 42 (42/0) | 45.0 (10.0)<br>51.0 (9.7)<br>53.1 (12.3) | 40.0 (14.9) | LCAT | Serum | Spectro. | 6 | 7,5 | LCAT * |
| 160 | (Chung et al. 2013) | USA | Depression | HC | 18 (6/12) | 36 (12/24) | 54 (18/36) | 32.2 (10.0) | 32.3 (7.6) | 8-Isoprostane | Urine | GC/MS | 4 | 13 | 8-Isoprostane # |
| 161 | (Kalenderolu et al. 2010) | Turkey | BD | HC | 60 (-/-) | 30 (-/-) | 90 (-/-) | 34.38 (11.05) | - | Ghrelin, HDL | Serum | ELISA, Spectro. | 5 | 11 | Ghrelin * , HDL |
| 162 | (Jirakran, Vasupanrajit et al. 2023) | Thailand | MDD | HC | 66 (18/48) | 67 (9/58) | 133 (27/106) | 36.9 (11.5) | 37.9 (9.2) | Apo A, Apo B, HDL, LCAT, | Serum | Spectro. | 6 | 6 | Apo A*, Apo B #, HDL*, LCAT* |
| 163 | (Ekinci and Ekinci 2017) | Turkey | Depression | HC | 37 (15/22)<br>120 (27/75) | 50 (13/37) | 87 (28/59)<br>170 (40/112) | 43 (14.1)<br>41.8 (11.4) | 44.12(4.2) | HDL | Serum | Automated | 6 | 9 | HDL * |
| 164 | (Eidan, Al-Harmoosh et al. 2019) | Iraq | Depression | HC | 22 (16/6) | 30 (13/17) | 52 (29/23) | 36.91(10.3) | 31.1(15.4) | HDL | Plasma | Spectro. | 3 | 15 | HDL # |
| 165 | (Maes, Simeonova et al. 2019) | Belgium | MDD | HC | 54 (23/31) | 37 (12/25) | 91 (35/56) | 43.5 (11.6) | 43.6 (11.1) | oxLDL, Peroxide | Serum | ELISA | 7 | 9 | oxLDL #, Peroxide # |
| 166 | (Simeonova, Stoyanov et al. 2020a) | Belgium | MDD<br>BD | HC | 44 (23/21)<br>37 (13/24) | 30 (10/20) | 74 (33/41)<br>67 (23/44) | 45.6 (12.3)<br>42.2 (12.4)<br>39.1 (12.8) | 38.9 (11.7) | IgG-oxLDL, Peroxides | Serum | ELISA, Spectro. | 8 | 6,5 | IgG-oxLDL#, peroxides # |
| 167 | (Ogłodek 2017) | Poland | MDD | HC | 60 | 40 | 100 | 45.2 (4.5) | 0 | PON1 | Serum | ELISA | 5 | 11,5 | PON1* |

| | | | | | | | | | | | | | | | |
|---|---|---|---|---|---|---|---|---|---|---|---|---|---|---|---|
| | | | MDD+PTSD | | (30/30) | (20/20) | (50/50) | | | | | | | | |
| 168 | (Simeonova, Stoyanov et al. 2021b) | Belgium | MDD | HC | 47 (24/23) | 35 (14/21) | 82 (38/44) | 45.5 (12.2) | 41.8 (12.8) | IgG-oxLDL, IgM-MDA, Peroxides | Serum | ELISA, Spectro. | 5 | 10 | IgG-oxLDL #, IgM-MDA #, peroxides # |
| | | | BD | | 29 (12/17) | | 64 (26/38) | 42.6 (12.6) | | | | | | | |
| | | | | | 25 (10/15) | | 60 (24/36) | 37.2 (13.7) | | | | | | | |
| 169 | (Maes, Kubera et al. 2013b) | Belgium | MDD | HC | 74 (36/38) | 57 (13/44) | 131 (49/82) | 40.3 (9.8) | 40.3 (9.8) | IgM-MDA | Serum | ELISA | 6 | 8,5 | IgM-MDA # |

*: Indicates that patients have reduced level of the measured metabolite compared to healthy control

#: Indicates that patients have increased level of the measured metabolites compared to healthy control

MDD: Major depression disorder, BD: Bipolar disorder, MDE: Major depressive episode, UP: Unipolar depression, TRD: Treatment resistant depression, PPD: post-partum depression, Apo A: Apolipoprotein A, Apo E: Apolipoprotein E, 4-HNE: 4-Hydroxynonenal, MDA: Malonaldehyde, PON-1: Paraoxonase-1, HDL: High-density lipoprotein, LCAT: Lecithin–cholesterol acyltransferase, ND: No difference, HC: Healthy control, Spectro.: Spectrophotometer.

**ESF, Table 6. The secondary outcomes (single indicators) and number of patients with affective disorders and healthy controls and the side of standardized mean differences (SMD) and the 95% confidence intervals with respect to the zero SMD.**

| Outcome single indicators | n studies | Side of 95% confidence intervals | | | | Patient Cases | Control Cases | Total number of participants |
|---|---|---|---|---|---|---|---|---|
| | | < 0 | Overlap 0 and SMD < 0 | Overlap 0 and SMD > 0 | > 0 | | | |
| HDL | 53 | 18 | 18 | 11 | 6 | 10326 | 11027 | 21353 |
| PON1 | 8 | 2 | 5 | 1 | 0 | 413 | 580 | 993 |
| ApoA | 9 | 5 | 1 | 2 | 1 | 583 | 551 | 1134 |
| ApoE | 3 | 1 | 0 | 2 | 0 | 50 | 76 | 126 |
| Vitamin A | 3 | 1 | 2 | 0 | 0 | 180 | 147 | 327 |
| Vitamin D | 15 | 10 | 3 | 2 | 0 | 1878 | 1367 | 3245 |
| Vitamin E | 9 | 4 | 2 | 2 | 1 | 629 | 553 | 1182 |
| Ghrelin | 20 | 5 | 2 | 4 | 9 | 630 | 681 | 1311 |
| MDA | 63 | 3 | 7 | 12 | 41 | 3061 | 2991 | 6052 |
| Isoprostane | 9 | 0 | 1 | 2 | 6 | 352 | 326 | 678 |
| 4-HNE | 9 | 0 | 2 | 4 | 3 | 279 | 246 | 525 |
| Peroxides | 8 | 0 | 1 | 1 | 6 | 472 | 235 | 707 |
| LOOH | 8 | 1 | 2 | 3 | 2 | 399 | 346 | 745 |
| IgM-MDA | 4 | 0 | 0 | 1 | 3 | 221 | 148 | 369 |
| IgG-oxLDL | 5 | 0 | 0 | 0 | 5 | 247 | 152 | 399 |
| OxLDL | 4 | 1 | 0 | 0 | 3 | 244 | 125 | 369 |

HDL: high-density lipoprotein cholesterol, PON1: paraoxonase 1, Apo: apolipoprotein, LCAT: lecithin cholesterol acyltransferase, MDA: malondialdehyde, HNE: hydroxynonenal, LOOH: lipid hydroperoxides, oxLDL: oxidized low-density lipoprotein cholesterol

**ESF, Table 7. Results of meta-analysis performed on several secondary outcome variables.**

| Outcome single indicators | n | Groups | SMD | 95% CI | z | p | Q | df | p | $I^2$ (%) | $\tau^2$ | T |
|---|---|---|---|---|---|---|---|---|---|---|---|---|
| HDL* | 53 | Overall | -0.227 | -0.347; -0.108 | -3.723 | <0.0001 | 536.916 | 52 | <0.0001 | 90.315 | 0.154 | 0.393 |
| PON1* | 8 | Overall | -0.416 | -0.761; -0.072 | -2.368 | 0.018 | 39.264 | 7 | <0.0001 | 82.172 | 0.189 | 0.435 |
| ApoA* | 9 | Overall | -0.287 | -0.622; 0.047 | -1.683 | 0.092 | 54.689 | 8 | <0.0001 | 85.372 | 0.210 | 0.459 |
| ApoE* | 3 | Overall | -0.188 | -0.922; 0.546 | -0.501 | 0.616 | 6.192 | 2 | 0.045 | 67.702 | 0.283 | 0.532 |
| Vitamin A* | 3 | Overall | -0.320 | -0.540; -0.099 | -2.842 | 0.004 | 1.866 | 2 | 0.393 | 0.000 | 0.000 | 0.000 |
| Vitamin D* | 15 | Overall | -0.558 | -0.826; -0.291 | -4.090 | <0.0001 | 142.853 | 14 | <0.0001 | 90.200 | 0.228 | 0.478 |
| Vitamin E* | 9 | Overall | -0.476 | -1.082; -0.131 | -1.537 | 0.124 | 170.486 | 8 | <0.0001 | 95.308 | 0.807 | 0.899 |
| Ghrelin | 7 | BD | -0.449 | -1.325; 0.427 | -1.005 | 0.315 | 120.804 | 6 | <0.0001 | 95.033 | 1.321 | 1.149 |
| | 13 | Depression | 0.622 | 0.043; 1.200 | 2.107 | 0.035 | 164.827 | 12 | <0.0001 | 92.720 | 1.033 | 1.016 |
| | **BD vs MDD Q=3.998, df=1, p= 0.046** | | | | | | | | | | | |
| MDA | 27 | BD | 1.117 | 0.811; 1.423 | 7.152 | <0.0001 | 282.140 | 26 | <0.0001 | 90.785 | 0.577 | 0.760 |
| | 36 | Depression | 0.687 | 0.395; 0.979 | 4.609 | <0.0001 | 565.451 | 36 | 0.000 | 93.810 | 0.727 | 0.852 |
| | **BD vs MDD Q=3.961, df=1, p= 0.047** | | | | | | | | | | | |
| 8- Isoprostane* | 9 | Overall | 0.670 | 0.386;0.954 | 4.631 | <0.0001 | 23.754 | 8 | 0.003 | 66.321 | 0.118 | 0.344 |
| 4-HNE* | 9 | Overall | 0.376 | 0.040;0.713 | 2.190 | 0.029 | 25.534 | 8 | 0.001 | 68.669 | 0.173 | 0.416 |
| Peroxides* | 8 | Overall | 0.524 | 0.260; 0.787 | 3.890 | <0.0001 | 17.486 | 7 | 0.015 | 59.968 | 0.087 | 0.294 |
| LOOH | 8 | Overall | 0.006 | -0.266; 0.277 | 0.042 | 0.967 | 20.179 | 7 | 0.005 | 65.311 | 0.092 | 0.303 |
| IgG-oxLDL* | 5 | Overall | 1.085 | 0.860; 1.311 | 9.433 | <0.0001 | 3.312 | 4 | 0.507 | 0.000 | 0.000 | 0.000 |
| IgM-MDA* | 4 | Overall | 0.705 | 0.485; 0.926 | 6.265 | <0.0001 | 1.809 | 3 | 0.613 | 0.000 | 0.000 | 0.000 |
| OxLDL* | 4 | Overall | 0.983 | -0.085; 2.051 | 1.804 | 0.071 | 56.874 | 3 | <0.0001 | 94.725 | 1.114 | 1.055 |

* No significant difference between bipolar disorder (BD) and major depression (MDD)
HDL: high-density lipoprotein cholesterol, PON1: paraoxonase 1, Apo: apolipoprotein, LCAT: lecithin cholesterol acyltransferase, MDA: malondialdehyde, HNE: hydroxynonenal, LOOH: lipid hydroperoxides, oxLDL: oxidized low-density lipoprotein cholesterol

**ESF, Table 8. Results on publication bias.**

| Outcome feature sets | Fail safe n | Z Kendall's τ | p | Egger's t test (df) | p | Missing studies (side) | After Adjusting |
|---|---|---|---|---|---|---|---|
| HDL | -10.597 | 1.695 | 0.045 | 1.439 (51) | 0.078 | 12 (Left) | -0.440 (-0.569; -0.310) |
| PON1 | -4.845 | 1.113 | 0.132 | 1.535(6) | 0.087 | 3 (Left) | -0.685 (-1.066; -0.304) |
| ApoA | -4.262 | 0.104 | 0.456 | 0.045 (7) | 0.482 | 0 | - |
| ApoE | -0.774 | 0.000 | 0.500 | 0.590(1) | 0.330 | 0 | |
| Vitamin A | -2.837 | 0.000 | 0.500 | 0.023(1) | 0.492 | 0 | |
| Vitamin D | -12.025 | 0.197 | 0.421 | 1.112(13) | 0.143 | 0 | |
| Vitamin E | -5.096 | 2.189 | 0.014 | 1.296 (7) | 0.117 | 0 | |
| Ghrelin (BD) | -4.665 | 0 | 0.500 | 0.023(5) | 0.491 | 0 | |
| Ghrelin (Depression) | 6.348 | 1.891 | 0.029 | 1.658 (11) | 0.062 | 3 (Right) | 1.071(0.374;1.768) |
| MDA (BD) | 21.052 | 2.168 | 0.015 | 2.646 (25) | 0.006 | 2 (Right) | 1.239 (0.890; 1.589) |
| MDA (Depression) | 15.952 | 1.920 | 0.027 | 1.113 (34) | 0.136 | 0 | |
| 8-Isoprostane | 7.964 | 0.104 | 0.458 | 0.525(7) | 0.307 | 0 | 0 |
| 4-HNE | 3.752 | 0.521 | 0.301 | 0.998(7) | 0.175 | 2 (Left) | 0.199 (-0.155; 0.553) |
| Peroxides | 6.172 | 0.618 | 0.268 | 0.813(6) | 0.223 | 2 (Left) | 0.450 (0.219; 0.681) |
| LOOH | 0.249 | 2.350 | 0.009 | 1.721 (6) | 0.068 | 0 | 0 |
| IgG-oxLDL | 9.399 | 0.244 | 0.403 | 0.582(3) | 0.300 | 0 | 0 |
| IgM-MDA | 6.051 | 1.698 | 0.044 | 1.770 (2) | 0.109 | 0 | 0 |
| OxLDL | 6.640 | 1.019 | 0.154 | 1.429(2) | 0.144 | 0 | 0 |

HDL: high-density lipoprotein cholesterol, PON1: paraoxonase 1, Apo: apolipoprotein, LCAT: lecithin cholesterol acyltransferase, MDA: malondialdehyde, HNE: hydroxynonenal, LOOH: lipid hydroperoxides, oxLDL: oxidized low-density lipoprotein cholesterol

**ESF, Table 9. Results of meta-regression**

| Variables | No. of Studies | Covariates | 1-sided p-value | Z-Value |
|---|---|---|---|---|
| RCT | 55 | Latitude | 0.0011 | -3.07 |
| | 22 | DOI | 0.033 | -1.84 |
| HDL | 53 | Latitude | 0.019 | -2.06 |
| | 7 | DOI | 0.025 | -1.95 |
| Ghrelin | 15 | Female | 0.011 | -2.29 |
| | 16 | Sample size | 0.013 | -2.20 |
| | 20 | BD vs UP | 0.019 | 2.071 |
| | 19 | Smoking free | 0.002 | 2.83 |
| LPANTIOX | 99 | Latitude | 0.0011 | -3.05 |
| | 94 | Smoking Free | 0.0093 | 2.35 |
| | 79 | Age | 0.0038 | -2.67 |
| Vitamin E | 4 | BMI | 0.024 | 1.96 |
| LPOSTOX | 13 | YMRS | 0.024 | 1.96 |
| MDA | 63 | BD or UP | 0.024 | -1.97 |
| | 10 | YMRS | 0.042 | 1.72 |
| | 24 | BMI | 0.016 | -2.14 |
| 8-Isoprostane | 5 | BMI | 0.003 | 2.72 |
| | 9 | Age | 0.026 | 1.93 |
| | | Plasma | 0.023 | -1.99 |
| | | Central-Peripheral | 0.001 | -2.94 |
| 4-HNE | 6 | Serum | 0.003 | -2.74 |
| | | Plasma | 0.034 | -1.82 |
| | 9 | BD vs UP | 0.036 | 1.79 |
| Peroxides | 8 | Latitude | 0.000 | 3.97 |
| | 4 | BMI | 0.0001 | 3.81 |
| | 6 | Age | 0.0000 | 3.91 |
| LPAUTO | 5 | Sample size | 0.028 | -1.91 |
| | | Female cases | 0.038 | -1.77 |
| (LPOSTOX+LPAUTO)/ LPANTIOX ratio | 176 | BD vs UP | 0.014 | -2.18 |
| | 16 | YMRS | 0.018 | 2.09 |
| | 139 | Age | 0.0006 | 3.24 |

HDL: high density lipoprotein cholesterol, MDA: malondialdehyde, HNE: hydroxynonenal.

RCT or reverse cholesterol transport comprises high-density lipoprotein (HDL), paraoxonase-1 (PON-1), apolipoprotein A (ApoA), lecithin cholesterol acyltransferase (LCAT), ApoE and ghrelin.
LPANTIOX or lipid-associated antioxidant defenses comprises RCT and ADECK.

LPOSTOX or lipid oxidative stress toxicity comprises MDA, TBARS, 8-Isoprostane, 4HNE, oxLDL, 7-Ketocholesterol, 27-Hydroxycholesterol, 25-Hydroxycholesterol, 24-Hydroxycholesterol, peroxides, LOOH and ApoB.
LPAUTO or autoimmunity against lipid associated neoepitopes. comprises immunoglobulin G against oxLDL and Immunoglobulin M against MDA.

#### HDL

| Study name | Year | Std diff in means | p-Value | Patients | Healthy Control |
|---|---|---|---|---|---|
| Olusi | 1996 | 0,883 | 0,000 | 100 | 100 |
| Maes-e | 1997 | -1,085 | 0,000 | 36 | 28 |
| Sevincok | 2001 | -0,932 | 0,002 | 27 | 24 |
| Ranjekar | 2003 | -0,104 | 0,775 | 10 | 31 |
| Huang-c | 2003 | 0,193 | 0,339 | 68 | 39 |
| Karlovic | 2004 | -0,310 | 0,176 | 38 | 39 |
| Huang-d | 2004 | 0,192 | 0,309 | 103 | 39 |
| Roohafza | 2005 | -0,420 | 0,142 | 25 | 25 |
| Huang-b | 2005 | -0,176 | 0,276 | 109 | 59 |
| Sarandol-b | 2006 | 0,556 | 0,006 | 86 | 36 |
| Lehto | 2008 | -0,460 | 0,011 | 63 | 61 |
| Barim | 2009 | -0,078 | 0,791 | 24 | 22 |
| Sagud-D | 2009 | -1,553 | 0,000 | 34 | 50 |
| Sagud-B | 2009 | -1,147 | 0,000 | 21 | 50 |
| Kodydkova | 2009 | -0,691 | 0,005 | 35 | 35 |
| Lehto-b | 2010 | -0,364 | 0,052 | 44 | 88 |
| Dortland | 2010 | -0,047 | 0,378 | 864 | 629 |
| Sadeghi | 2011 | -1,110 | 0,000 | 153 | 147 |
| Kotan | 2011 | -0,174 | 0,401 | 50 | 44 |
| Sukreet | 2011 | 0,001 | 0,997 | 100 | 52 |
| Hocaoglu | 2012 | 0,943 | 0,001 | 30 | 30 |
| Algul-a | 2013 | -2,133 | 0,000 | 20 | 40 |
| Vargas-b-B | 2014 | -0,385 | 0,025 | 49 | 119 |
| Bortolasci-B | 2014 | -0,366 | 0,006 | 82 | 199 |
| Bortolasci-D | 2014 | -0,215 | 0,090 | 91 | 199 |
| Vargas-b-D | 2014 | -0,223 | 0,109 | 92 | 119 |
| Patra | 2014 | 0,058 | 0,823 | 30 | 30 |
| Sarandol-a | 2015 | -0,276 | 0,401 | 15 | 25 |
| Kahl | 2015 | -0,259 | 0,438 | 10 | 90 |
| Carmo | 2015 | 1,723 | 0,000 | 22 | 27 |
| Ekinci | 2017 | -1,681 | 0,000 | 70 | 50 |
| Tuncel-a | 2017 | -1,956 | 0,000 | 30 | 30 |
| Messaoud | 2017 | -0,344 | 0,018 | 81 | 151 |
| Moreira | 2017 | -0,536 | 0,000 | 55 | 863 |
| Baghai | 2018 | -0,416 | 0,003 | 100 | 104 |
| Pilla | 2018 | 0,390 | 0,000 | 186 | 250 |
| Enko | 2018 | 0,408 | 0,002 | 94 | 152 |
| Mendoza | 2018 | -0,168 | 0,090 | 204 | 206 |
| Hu | 2019 | 0,145 | 0,308 | 99 | 99 |
| Wagner | 2019 | -0,218 | 0,162 | 130 | 61 |
| Hui | 2019 | -0,038 | 0,870 | 37 | 37 |
| Su-a-B | 2019 | 0,035 | 0,814 | 92 | 89 |
| Su-b-D | 2019 | 0,136 | 0,288 | 195 | 89 |
| Eidan | 2019 | 0,372 | 0,189 | 22 | 30 |
| Karadeniz | 2020 | -0,046 | 0,849 | 37 | 33 |
| Mehrabani | 2021 | 0,076 | 0,593 | 75 | 150 |
| Guidara | 2021 | 0,383 | 0,123 | 33 | 33 |
| Kennedy | 2021 | -0,099 | 0,618 | 55 | 47 |
| Wei | 2022 | -0,401 | 0,000 | 6005 | 5810 |
| Kasak | 2022 | -0,375 | 0,108 | 39 | 36 |
| Aslan | 2022 | -0,264 | 0,042 | 134 | 108 |
| Shapiro | 2022 | 0,026 | 0,861 | 88 | 89 |
| Jirakran | 2023 | -0,532 | 0,030 | 35 | 34 |
| Pooled | | -0,227 | 0,000 | 10326 | 11027 |
| Prediction Interval | | -0,227 | | | |

Statistics for each study; Sample size

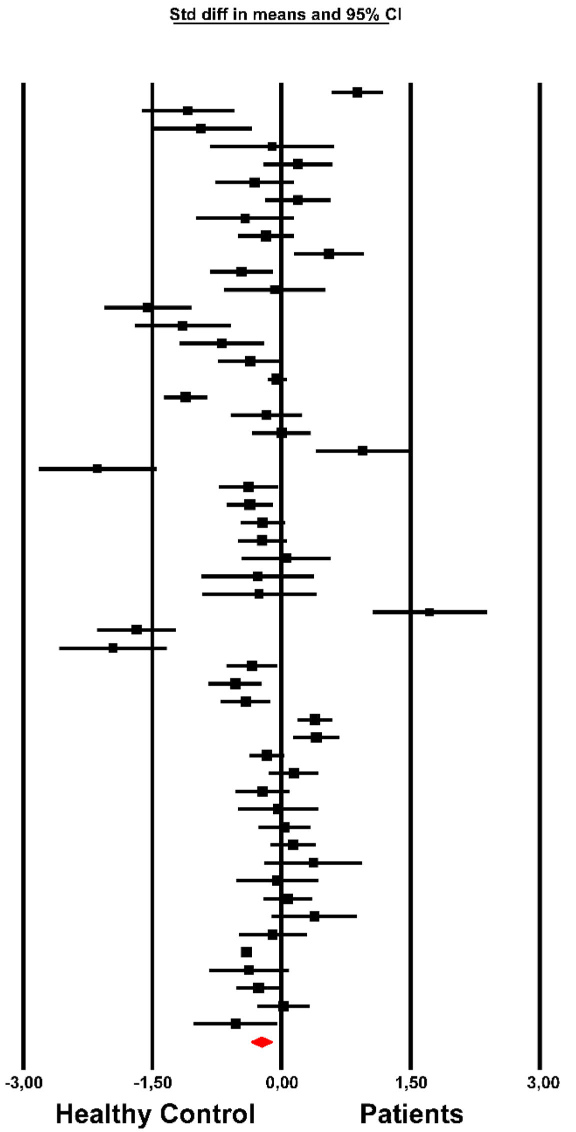

**Almulla et al, 2023**

**ESF, Figure 1.** The forest plot of high-density lipoprotein (HDL) in patients with major depression and bipolar disorder compared to healthy controls.

#### PON-1

| Study name | Year | Statistics for each study: Std diff in means | p-Value | Sample size: Patients | Healthy Control |
|---|---|---|---|---|---|
| Sarandol-b | 2006 | -0,094 | 0,638 | 86 | 36 |
| Barim | 2009 | -0,549 | 0,068 | 24 | 22 |
| Kodydkova | 2009 | -0,224 | 0,351 | 35 | 35 |
| Kotan | 2011 | -0,189 | 0,361 | 50 | 44 |
| Bortolasci-B | 2014 | -0,143 | 0,277 | 82 | 199 |
| Bortolasci-D | 2014 | -0,251 | 0,048 | 91 | 199 |
| Sarandol-a | 2015 | 0,048 | 0,884 | 15 | 25 |
| Oglodek | 2017 | -2,633 | 0,000 | 30 | 20 |
| Pooled | | -0,416 | 0,018 | 413 | 580 |
| Prediction Interval | | -0,416 | | | |

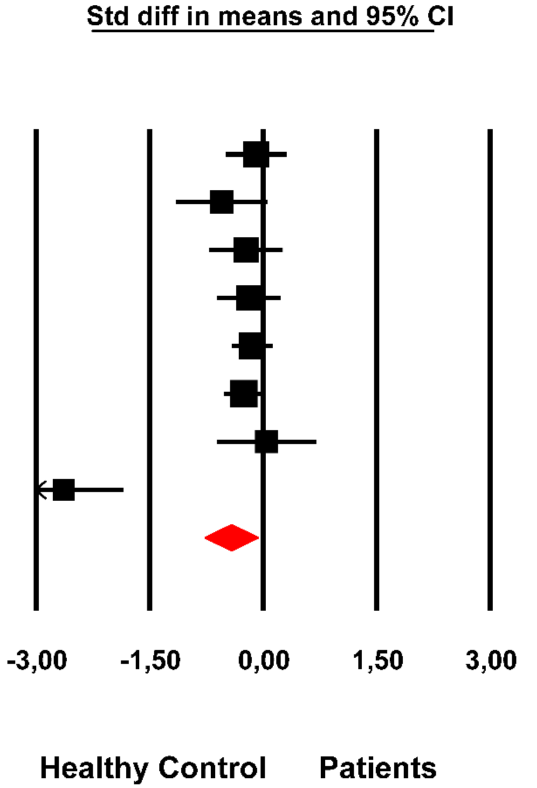

**Almulla et al, 2023**

**ESF, Figure 2**. The forest plot of paraoxonase 1 (PON-1) in patients with major depression and bipolar disorder compared to healthy controls.

**Vitamin A**

| Study name | Year | Statistics for each study: Std diff in means | p-Value | Sample size: Patients | Healthy Control |
|---|---|---|---|---|---|
| Kotan | 2011 | -0,217 | 0,296 | 50 | 44 |
| Chowdhury | 2017 | -0,536 | 0,006 | 55 | 55 |
| XUE | 2020 | -0,206 | 0,267 | 75 | 48 |
| Pooled | | -0,320 | 0,004 | 180 | 147 |
| Prediction Interval | | | | | |

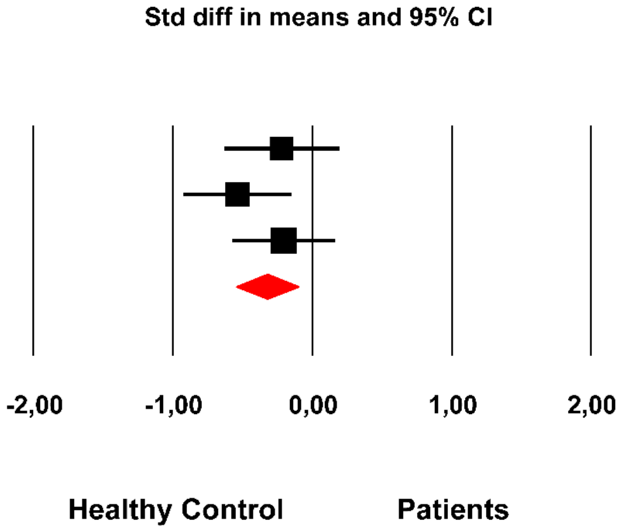

**Almulla et al, 2023**

**ESF, Figure 3.** The forest plot of vitamin A in patients with major depression and bipolar disorder compared to healthy controls.

#### Vitamin D

| Study name | Year | Statistics for each study: Std diff in means | p-Value | Sample size: Patients | Healthy Control |
|---|---|---|---|---|---|
| Schneider | 2000 | -0,774 | 0,005 | 25 | 31 |
| Jamilian | 2013 | -0,628 | 0,000 | 100 | 100 |
| Milaneschi | 2013 | -0,238 | 0,000 | 946 | 494 |
| Olga | 2014 | -1,755 | 0,000 | 91 | 89 |
| Abedi | 2018 | -0,619 | 0,001 | 60 | 60 |
| Altunsoy | 2018 | -0,669 | 0,008 | 29 | 40 |
| Petrov-B | 2018 | 0,665 | 0,106 | 13 | 12 |
| Petrov-D | 2018 | -0,120 | 0,775 | 11 | 12 |
| Grudet | 2020 | -0,185 | 0,352 | 48 | 54 |
| Okasha-a | 2020 | -1,041 | 0,002 | 20 | 20 |
| Esnafoglu | 2020 | -1,620 | 0,000 | 89 | 43 |
| Kamalzadeh | 2021 | -0,499 | 0,000 | 174 | 173 |
| Quessada | 2021 | -0,048 | 0,713 | 117 | 117 |
| khan | 2022 | -0,787 | 0,000 | 100 | 100 |
| Rheenen | 2023 | 0,363 | 0,152 | 55 | 22 |
| Pooled | | -0,558 | 0,000 | 1878 | 1367 |
| Prediction Interval | | -0,558 | | | |

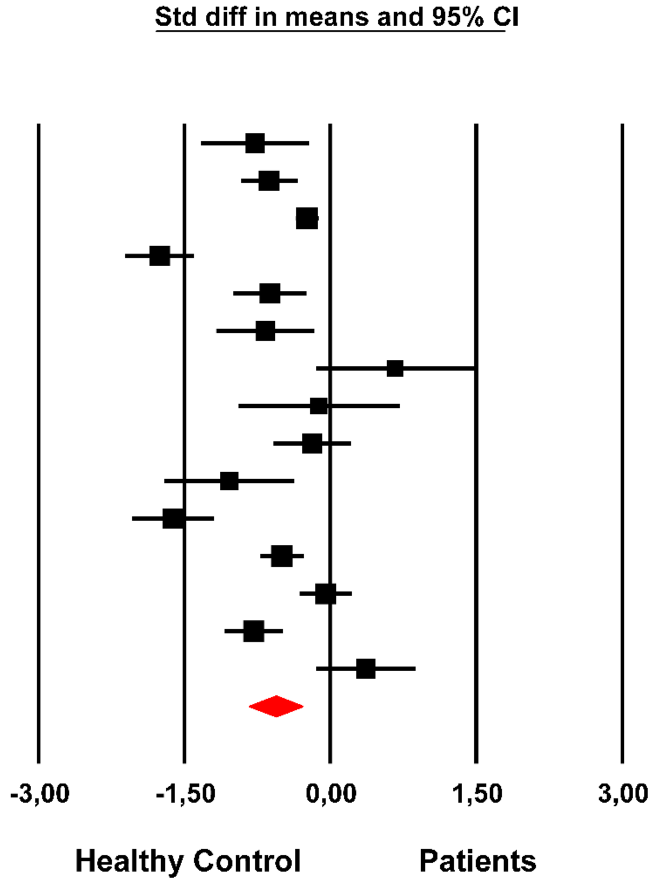

**Almulla et al, 2023**

**ESF, Figure 4**. The forest plot of vitamin D in patients with major depression and bipolar disorder compared to healthy controls.

### MDA

| Study name | Year | Subgroup within study | Std diff in means | p-Value | Sample size: Patients | Sample size: Healthy Control |
|---|---|---|---|---|---|---|
| Kuloglu | 2002 | BD | 3,236 | 0,000 | 23 | 20 |
| Ranjekar | 2003 | BD | 0,495 | 0,275 | 10 | 10 |
| Andreazza-b | 2007 | BD | 0,987 | 0,000 | 28 | 32 |
| Vieira | 2007 | BD | 2,217 | 0,000 | 23 | 30 |
| Kunz | 2008 | BD | 1,560 | 0,000 | 28 | 32 |
| Aksoy | 2010 | BD | 1,247 | 0,000 | 30 | 30 |
| Can-B | 2011 | BD | 2,203 | 0,000 | 39 | 30 |
| Kapczinski-B | 2011 | BD | 3,263 | 0,000 | 20 | 80 |
| Sukreet | 2011 | BD | 0,738 | 0,000 | 100 | 52 |
| Banerjee | 2012 | BD | 2,207 | 0,000 | 37 | 35 |
| Ezzaher | 2012 | BD | 0,640 | 0,000 | 90 | 92 |
| Magalhaes-B | 2012 | BD | 1,462 | 0,000 | 82 | 94 |
| Gubert | 2013 | BD | 0,488 | 0,239 | 12 | 12 |
| Aydemir | 2014 | BD | 1,122 | 0,000 | 51 | 50 |
| Cudney | 2014 | BD | 0,306 | 0,185 | 52 | 30 |
| Sousa | 2014 | BD | -0,363 | 0,207 | 29 | 22 |
| Bengesser | 2015 | BD | -0,224 | 0,131 | 113 | 78 |
| Sarandol-a | 2015 | BD | 0,389 | 0,242 | 15 | 25 |
| Tsai-a | 2015 | BD | 0,735 | 0,006 | 23 | 40 |
| Tuncel-c | 2015 | BD | 1,018 | 0,004 | 18 | 18 |
| Siwek | 2016 | BD | 0,826 | 0,000 | 43 | 50 |
| Akgun | 2017 | BD | 1,289 | 0,000 | 20 | 22 |
| Chowdhury | 2017 | BD | 0,545 | 0,005 | 55 | 55 |
| Ceylan | 2018 | BD | -0,017 | 0,939 | 32 | 51 |
| Kucma-a-B | 2018 | BD | 0,803 | 0,000 | 133 | 50 |
| Qinyu-b | 2019 | BD | 1,700 | 0,000 | 61 | 49 |
| Qinyu | 2020 | BD | 1,994 | 0,000 | 28 | 49 |
| Pooled | | BD | 1,117 | 0,000 | 1194 | 1138 |
| Prediction Interval | | BD | 1,117 | | | |
| Fadillioglu | 2000 | Depression | 0,103 | 0,855 | 8 | 8 |
| Bilici | 2001 | Depression | 1,367 | 0,000 | 15 | 32 |
| Ozcan | 2003 | Depression | 1,144 | 0,000 | 30 | 21 |
| Sarandol-b | 2006 | Depression | 1,546 | 0,000 | 86 | 36 |
| Sarandol-c | 2007 | Depression | 0,776 | 0,000 | 96 | 54 |
| Galecki | 2009 | Depression | 0,671 | 0,005 | 50 | 30 |
| Can-D | 2011 | Depression | 2,169 | 0,000 | 32 | 30 |
| Kapczinski-D | 2011 | Depression | 0,955 | 0,001 | 20 | 80 |
| Kotan | 2011 | Depression | 0,771 | 0,000 | 50 | 44 |
| Bal | 2012 | Depression | 0,901 | 0,000 | 42 | 38 |
| Ghodake | 2012 | Depression | 0,871 | 0,001 | 30 | 30 |
| Magalhaes-D | 2012 | Depression | -0,802 | 0,000 | 82 | 94 |
| Stefanescu | 2012 | Depression | 1,293 | 0,001 | 16 | 20 |
| Baek | 2013 | Depression | 0,296 | 0,063 | 80 | 80 |
| Rybka | 2013 | Depression | 1,635 | 0,000 | 15 | 19 |
| Vargas | 2013 | Depression | -0,079 | 0,615 | 70 | 101 |
| Bajpai | 2014 | Depression | 1,945 | 0,000 | 60 | 40 |
| Spanemberg | 2014 | Depression | -0,101 | 0,725 | 17 | 54 |
| Cimen | 2015 | Depression | 0,938 | 0,041 | 18 | 18 |
| Kaufmann | 2015 | Depression | 0,641 | 0,002 | 49 | 49 |
| Camkurt-a | 2016 | Depression | 3,353 | 0,000 | 50 | 50 |
| Tsai-b | 2016 | Depression | 0,483 | 0,077 | 21 | 40 |
| Camkurt-b | 2017 | Depression | -0,184 | 0,443 | 33 | 37 |
| Diniz | 2018 | Depression | 0,101 | 0,587 | 77 | 47 |
| Islam-a | 2018 | Depression | 1,436 | 0,000 | 247 | 248 |
| Jordan | 2018 | Depression | 0,325 | 0,249 | 18 | 43 |
| Kucma-a-D | 2018 | Depression | 0,842 | 0,000 | 114 | 50 |
| Kucma-b | 2018 | Depression | 0,939 | 0,000 | 57 | 50 |
| Gaytan | 2019 | Depression | -0,313 | 0,451 | 12 | 12 |
| Maes | 2019 | Depression | 0,058 | 0,727 | 105 | 54 |
| Alamolhoda | 2020 | Depression | 0,401 | 0,112 | 22 | 58 |
| Hamed | 2020 | Depression | -0,647 | 0,001 | 60 | 60 |
| Mehrabani | 2021 | Depression | -1,080 | 0,000 | 75 | 150 |
| Alvarez-Mon | 2022 | Depression | 0,829 | 0,006 | 30 | 20 |
| Miniksar | 2022 | Depression | 1,123 | 0,000 | 42 | 38 |
| Opankovic | 2022 | Depression | 0,431 | 0,134 | 39 | 18 |
| Pooled | | Depression | 0,687 | 0,000 | 1867 | 1853 |
| Prediction Interval | | Depression | 0,687 | | | |
| Pooled | | Overall | 0,892 | 0,000 | 3061 | 2991 |
| Prediction Interval | | Overall | 0,892 | | | |

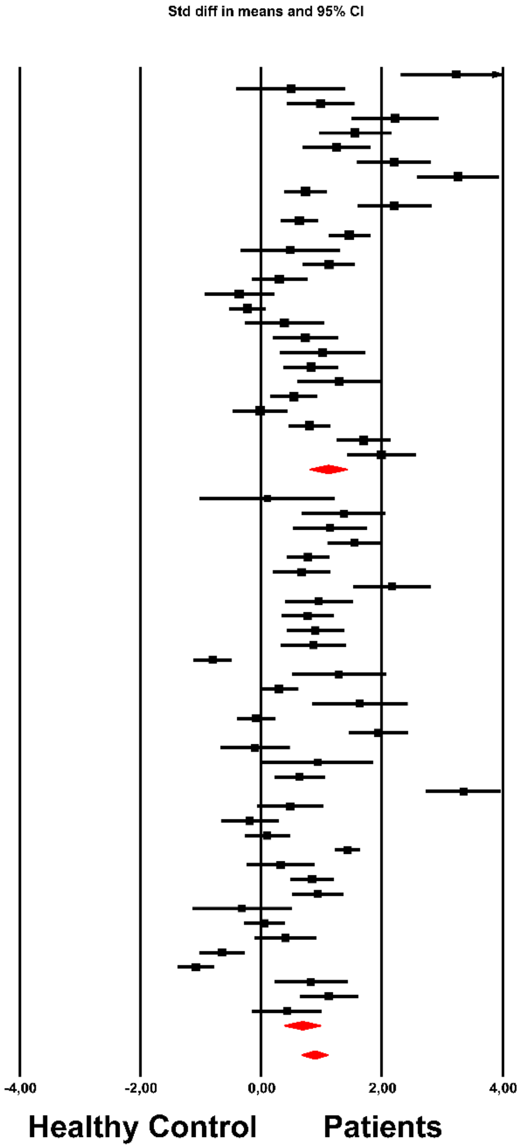

Almulla et al, 2023

**ESF, Figure 5**. The forest plot of malonaldehyde (MDA) in patients with major depression and bipolar disorder (BD) compared to healthy controls.

#### Isoprostane

| Study name | Year | Statistics for each study: Std diff in means | p-Value | Sample size: Patients | Healthy Control |
|---|---|---|---|---|---|
| Andreazza-c | 2013 | 1,227 | 0,000 | 16 | 26 |
| Chopra | 2021 | 0,521 | 0,019 | 42 | 42 |
| Diniz | 2018 | 0,546 | 0,004 | 77 | 47 |
| Lindqvist | 2017 | 0,375 | 0,061 | 47 | 55 |
| Rawdin | 2013 | -0,064 | 0,841 | 20 | 19 |
| Scolaa | 2015 | 0,110 | 0,769 | 16 | 13 |
| Yager | 2010 | 0,706 | 0,000 | 73 | 72 |
| Pooled | | 0,510 | 0,000 | 291 | 274 |
| Prediction Interval | | 0,510 | | | |

Std diff in means and 95% CI

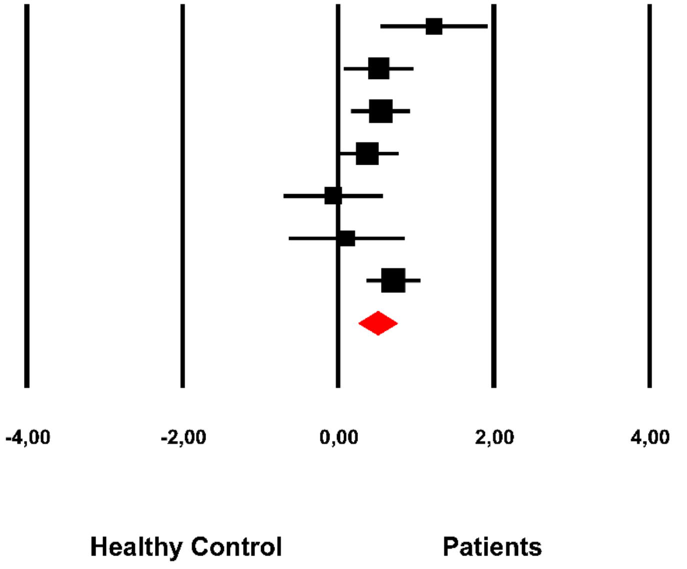

**Almulla et al, 2023**

**ESF, Figure 6**. The forest plot of isoprostane in patients with major depression and bipolar disorder compared to healthy controls.

#### 4-HNE

| Study name | Year | Std diff in means | p-Value | Patients | Healthy Control |
|---|---|---|---|---|---|
| Selley | 2004 | 1,223 | 0,000 | 25 | 25 |
| Wang-D | 2009 | 0,528 | 0,155 | 15 | 15 |
| Wang-B | 2009 | 0,895 | 0,019 | 15 | 15 |
| Andreazza-c | 2013 | 0,583 | 0,072 | 16 | 26 |
| Versace | 2014 | -0,366 | 0,237 | 24 | 19 |
| Andreazza-a | 2015 | 0,121 | 0,418 | 110 | 75 |
| Scolaa | 2015 | 0,255 | 0,497 | 16 | 13 |
| Elvsashagen | 2016 | 0,670 | 0,010 | 29 | 33 |
| Newton | 2017 | -0,311 | 0,257 | 29 | 25 |
| Pooled | | 0,376 | 0,029 | 279 | 246 |
| Prediction Interval | | 0,376 | | | |

Statistics for each study; Sample size

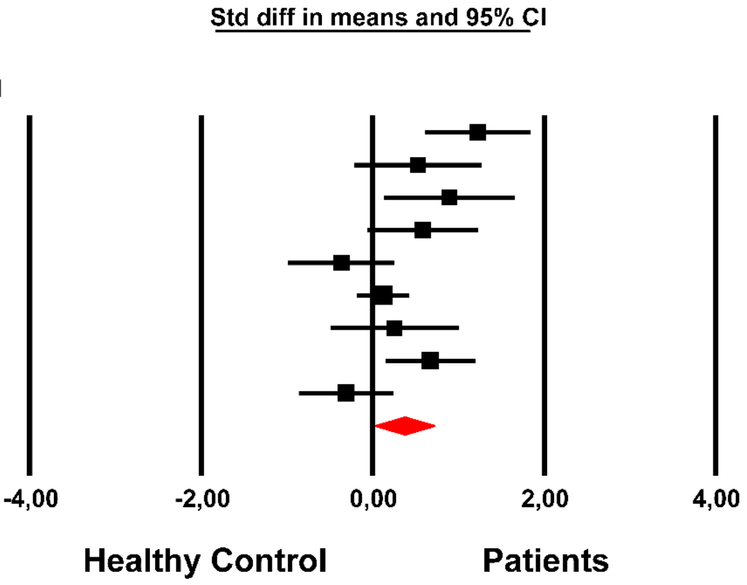

**Almulla et al, 2023**

**ESF, Figure 7**. The forest plot of 4-hydroxynonenal (4-HNE) in patients with major depression and bipolar disorder compared to healthy controls.

#### Peroxides

| Study name | Year | Std diff in means | p-Value | Patients | Healthy Control |
|---|---|---|---|---|---|
| Maes-b | 2010 | 0,409 | 0,062 | 54 | 35 |
| Maes-h | 2013 | 0,718 | 0,001 | 113 | 28 |
| Maes-k | 2019 | 0,631 | 0,009 | 96 | 22 |
| Simeon-b-BD | 2020 | 0,758 | 0,004 | 33 | 30 |
| Simeon-b-D | 2020 | 0,720 | 0,003 | 44 | 30 |
| Simeon-BD | 2021 | 0,610 | 0,020 | 27 | 35 |
| Simeon-D | 2021 | 0,735 | 0,001 | 47 | 35 |
| Katrencikova | 2021 | -0,449 | 0,086 | 58 | 20 |
| Pooled | | 0,524 | 0,000 | 472 | 235 |
| Prediction Interval | | 0,524 | | | |

(Column groups: Statistics for each study — Std diff in means, p-Value; Sample size — Patients, Healthy Control)

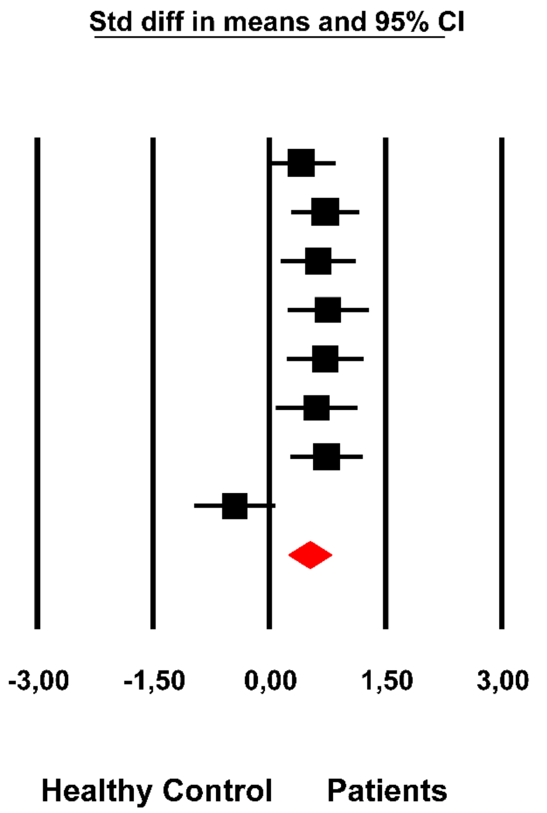

Almulla et al, 2023

**ESF, Figure 8.** The forest plot of peroxides in patients with major depression and bipolar disorder compared to healthy controls.

#### ESF, Listing of eligible studies

(Maes, Delanghe et al. 1994, Olusi and Fido 1996, Maes, Smith et al. 1997, Maes, Christophe et al. 1999, Fadillioğlu, Kaya et al. 2000, Maes, De Vos et al. 2000, Schneider, Weber et al. 2000, Bilici, Efe et al. 2001, Sevincok, Buyukozturk et al. 2001, Kuloglu, Ustundag et al. 2002, Huang, Wu et al. 2003, Khanzode, Dakhale et al. 2003, Ranjekar, Hinge et al. 2003, Huang and Chen 2004, Karlović, Buljan et al. 2004, Ozcan, Gulec et al. 2004, Selley 2004, Digney, Keriakous et al. 2005, Hamidreza Roohafza 2005, Huang 2005, Sarandol, Sarandol et al. 2006, Andreazza, Cassini et al. 2007, Eskandari, Martinez et al. 2007, Kurt, Guler et al. 2007, Machado-Vieira, Andreazza et al. 2007, Sarandol, Sarandol et al. 2007, Dean, Digney et al. 2008, Dimopoulos, Piperi et al. 2008, Kunz, Gama et al. 2008, Kurt, Güler et al. 2008, Lehto, Hintikka et al. 2008, Barim, Aydin et al. 2009, Gałecki, Szemraj et al. 2009, Kluge, Schussler et al. 2009, Kodydková, Vávrová et al. 2009, Maes, Mihaylova et al. 2009, Sagud, Mihaljevic-Peles et al. 2009, Wang, Shao et al. 2009, Kalenderoglu, Savafi et al. 2010, Lehto, Niskanen et al. 2010, Maes, Mihaylova et al. 2010, S. Nur Aksoy and Abdurrahman Altındag 2010, van Reedt Dortland, Giltay et al. 2010, Yager, Forlenza et al. 2010, Kapczinski, Dal-Pizzol et al. 2011, Kotan, Sarandol et al. 2011, Murat Can 2011, Sadeghi, Roohafza et al. 2011, Sonal Sukreet and Chaturvedi 2011, Vila-Rodriguez, Honer et al. 2011, Bal, Acar et al. 2012, Banerjee, Dasgupta et al. 2012, Ezzaher, Haj Mouhamed et al. 2012, Ghodake, Suryakar et al. 2012, Hocaoglu, Kural et al. 2012, Ishitobi, Kohno et al. 2012, Magalhães, Jansen et al. 2012, Matsuo, Nakano et al. 2012, Pomara, Bruno et al. 2012, Stefanescu and Ciobica 2012, Andreazza, Wang et al. 2013, Baek and Park 2013, Chung, Schmidt et al. 2013, Erman, Erman et al. 2013, Gubert, Stertz et al. 2013, Jamilian, Bagherzadeh et al. 2013, Maes, Kubera et al. 2013, Rawdin, Mellon et al. 2013, Rybka, Kędziora-Kornatowska et al. 2013, Vargas, Nunes et al. 2013, Vuksan-Cusa, Sagud et al. 2013, Maes, Kubera et al. 2013a, Aydemir, Çubukçuoğlu et al. 2014, Bajpai, Verma et al. 2014, Bortolasci, Vargas et al. 2014, Cudney, Sassi et al. 2014, de Sousa, Zarate et al. 2014, Józefowicz, Rabe-Jabłońska et al. 2014, Milaneschi, Hoogendijk et al. 2014, Ozsoy, Besirli et al. 2014, Patra, Khandelwal et al. 2014, Spanemberg, Caldieraro et al. 2014, Vargas, Nunes et al. 2014, Versace, Andreazza et al. 2014, Andreazza, Gildengers et al. 2015, Bengesser, Lackner et al. 2015, Cimen, Gumus et al. 2015, Kahl, Schweiger et al. 2015, Kaufmann, Gazal et al. 2015, Ormonde do Carmo, Mendes-Ribeiro et al. 2015, Rosso, Cattaneo et al. 2015, Sarandol, Sarandol et al. 2015, Tsai and Huang 2015, Tunçel, Sarısoy et al. 2015, Camkurt, Fındıklı et al. 2016, Elvsåshagen, Zuzarte et al. 2016, Mondin, de Azevedo Cardoso et al. 2016, Scola, McNamara et al. 2016, Siwek, Sowa-Kucma et al. 2016, Tsai and Huang 2016, Tunçel Ö, Akbaş et al. 2016, Akgün, Köken et al. 2017, Camkurt, Fındıklı et al. 2017, Chowdhury, Hasan et al. 2017, Ekinci and Ekinci 2017, Lindqvist, Dhabhar et al. 2017, Messaoud, Mensi et al. 2017, Moreira, Jansen et al. 2017, Newton, Naiberg et al. 2017, Ogłodek 2017, Abedi, Bovayri et al. 2018, Algul and Ozcelik 2018, Altunsoy, Yüksel et al. 2018, Baghai, Varallo-Bedarida et al. 2018, Ceylan, Tuna et al. 2018, Diniz, Mendes-Silva et al. 2018, Enko, Brandmayr et al. 2018, Islam, Islam et al. 2018, Jordan, Dobrowolny et al. 2018, Petrov, Aldoori et al. 2018, Ramachandran Pillai, Wilson et al. 2018, Segoviano-Mendoza, Cárdenas-de la Cruz et al. 2018, Sowa-Kućma, Styczeń et al. 2018, Sowa-Kućma, Styczeń et al. 2018, Tunçel Ö, Sarısoy et al. 2018, Baltazar-Gaytan, Aguilar-Alonso et al. 2019, Eidan, Al-Harmoosh et al. 2019, Hu, Wang et al. 2019, Hui, Yin et al. 2019, Lv, Guo et al. 2019, Maes, Landucci Bonifacio et al. 2019, Maes, Simeonova et al. 2019, Su, Li et al. 2019, Wagner, Musenbichler et al. 2019, Alamolhoda , Kariman et al. 2020, Esnafoglu and Ozturan 2020, Grudet, Wolkowitz et al. 2020, Hamed, Elmalt et al. 2020, Islam, Ali et al. 2020, Karadeniz, Yaman et al. 2020, Lv, Hu et al. 2020, Okasha, Sabry et al. 2020, Xue, Zeng et al. 2020, Simeonova, Stoyanov et al. 2020a, Chen, Hsu et al. 2021, Guidara, Messedi et al. 2021, Kamalzadeh, Saghafi et al. 2021, Katrenčíková, Vaváková et al. 2021, Kennedy, Islam et al. 2021, Parul Chopra 2021, Platzer, Fellendorf et al. 2021, Quessada, Nascimento et al. 2021, Vaghef-Mehrabani, Izadi et al. 2021, Simeonova, Stoyanov et al. 2021b, Abdel Aziz, Al-Mugaddam et al. 2022, Alvarez-Mon, Ortega et al. 2022, Draghici 2022, Huang, Chen et al. 2022, Kasak, Ceylan et al. 2022, khan, Shafiq et al. 2022, Kilicarslan, Sahan et al. 2022, Okasha, El-Gabry et al. 2022, Opanković, Milovanović et al. 2022, Shapiro, Kennedy et al. 2022, Ünler, Ekmekçi Ertek et al. 2022, Wei, Wang et al. 2022, Yıldız Miniksar and Göçmen 2022, Jirakran, Vasupanrajit et al. 2023, Van Rheenen, Ringin et al. 2023).
